# Comparative effectiveness of sulfadoxine–pyrimethamine plus amodiaquine versus other antimalarial regimens for paediatric malaria chemoprevention in the context of drug resistance: a systematic review and meta-analysis

**DOI:** 10.64898/2026.07.16.26356047

**Authors:** Gina Cuomo-Dannenburg, Andria Mousa, Oliver Simmons, Matt Cairns, Sarah Staedke, R Matthew Chico, Cally Roper, Patrick GT Walker, Lucy Okell

## Abstract

Each year, over 50 million children receive preventive malaria treatment. However, to date there has been no consensus on the most effective antimalarial drugs to use, especially given geographic differences in drug resistance. Here, we conduct a systematic review comparing the effectiveness of the most commonly used antimalarial chemopreventive regimen, sulfadoxine-pyrimethamine plus amodiaquine (SP+AQ), with other antimalarial drugs in preventing new infections. We searched MEDLINE, Embase, Global Health, PubMed and WWARN clinical trial databases until 06 December 2025 for studies satisfying the inclusion criteria. Studies were included if they were peer-reviewed, randomised-controlled studies in Africa, measuring incidence of infection or clinical episodes of *Plasmodium falciparum* malaria for at least 28 days post-treatment. We also compiled data on the prevalence of markers of resistance in the parasite *dhfr, dhps* and *mdr1* genes in the study areas. We conducted meta-analyses of incidence rates, with subgroup analyses by drug resistance levels. This review is registered on PROSPERO (CRD42024577149). We identified 27 studies representing 38,252 participants in 32 sites across 13 countries. In pooled analysis, SP+AQ reduced incidence of malaria by 54.6% (95% CI: 33.8–68.8%) compared to SP alone, including significantly outperforming SP even in areas with low SP resistance. These findings suggest that countries currently using SP alone for chemoprevention should consider switching to SP+AQ. Where AQ resistance remains low, available evidence suggests SP+AQ remains efficacious for malaria chemoprevention. SP+AQ was comparable to the artemisinin-based treatment, dihydroartemisinin-piperaquine across all studies (incidence rate ratio 0.93; 95% CI 0.78- 1.11). By resistance levels, SP+AQ had slightly higher efficacy in areas with low SP and AQ resistance but had comparable or slightly lower efficacy in areas with higher resistance. Using artemisinin-based treatments for chemoprevention must be balanced against the risk of worsening artemisinin resistance in Eastern and Southern Africa. This study was funded by the UK Royal Society.

## Background

Preventive antimalarial treatment (chemoprevention) is given to over 50 million young children each year to protect against *Plasmodium falciparum* malaria in areas with moderate to high transmission levels.^1^ The largest chemoprevention programmes in children are currently in the Sahel subregion of west and central Africa, which experiences intense but highly seasonal malaria transmission.^2^ Here, between three and five monthly cycles of sulfadoxine-pyrimethamine (SP) plus amodiaquine (AQ) are given to children aged under-five years during the transmission season as seasonal malaria chemoprevention (SMC). This intervention has achieved substantial reductions in clinical malaria episodes, severe disease and malaria-attributable mortality both in trials and in routine implementation evaluations.^3,4^ Regions in eastern and southern Africa also experience highly seasonal malaria transmission, but drug resistance to SP is widespread in these areas and, therefore, SMC was not originally deployed outside the Sahel.^2^ Other areas with less seasonal transmission have high malaria burden in children and might benefit from chemoprevention programmes. Perennial malaria chemoprevention (PMC) programmes, for example, have historically targeted younger children (< 1 year old) with three doses of SP alone at age 2, 3 and 9 months according to previous World Health Organization (WHO) guidelines, to be delivered alongside childhood vaccination schedules. More recently, 8 countries have implemented PMC programmes with many including children up to 2 years of age.^1^

In June 2022, WHO policy recommendations for malaria chemoprevention were updated.^5^ The new guidance was not prescriptive about the for antimalarial drugs to use, the number of doses and ages to target, and removed some previous restrictions related to geographic region and local resistance levels. Further evidence on choice of antimalarial regimen for chemoprevention outside the Sahel subregion is now needed, to confirm whether SP+AQ is optimal, or if other antimalarials are more effective in these areas. Mutations in the malaria parasites which confer partial drug resistance to SP and to AQ are widespread, but their prevalence varies considerably geographically and has changed over time. SP was formerly widely used as first-line treatment for malaria, but mutations in the *pfdhfr* and *pfdhps* genes (*dhfr* N51I + C59R + S108N and *dhps* A437G + K540E + A581G mutations)^6^ emerged and spread, with increasing numbers of mutations causing higher grade resistance. Due to rising treatment failure, countries switched to more efficacious artemisinin-combination therapies (ACTs), but SP continues to be used for chemoprevention. Parasites with higher-grade resistance to SP remain prevalent in parts of Eastern and Southern Africa, and chemoprevention efficacy of SP alone decreases as resistance increases ^7–11^. However, mutations conferring resistance to SP are less common in Western and Central Africa.^12^

AQ is predominantly used as a partner drug in both curative first-line malaria treatment, artesunate (AS) plus AQ, and as part of the SMC drug regimen SP+AQ. Mutations in the *pfcrt* and *pfmdr1* genes (*mdr1* N86Y, Y184, D124Y and *crt* K76T) have been associated with treatment failures^13,14^ and both *mdr1* N86Y and *crt* K76T with reduced prophylactic protection following AS+AQ.^15^ Prevalence of these mutations was historically high due to widespread use of chloroquine (CQ), a former first-line antimalarial drug related to amodiaquine. However, these mutations have declined considerably in frequency in recent years across most of the continent since CQ was withdrawn as the first-line treatment, potentially due to lower fitness and counter-selection by a key first-line treatment, artemether-lumefantrine.^16^

To date, there is limited evidence on chemoprevention efficacy with SP+AQ in areas of higher-grade SP resistance, with conflicting results. For example, one cluster randomised trial in Uganda found high efficacy against clinical disease, while another trial in Uganda, despite showing high efficacy against clinical disease, found limited efficacy against parasitaemia.^17–23^ Currently, the main widely considered alternative chemoprevention drug to SP or SP+AQ is the ACT dihydroartemisinin plus piperaquine (DHA+PPQ). DHA+PPQ has been evaluated across a variety of settings, including for intermittent preventive treatment in pregnancy (IPTp)^24^ and in schoolchildren^25,26^ and for post-discharge malaria chemoprevention (PDMC),^27^ but has not been adopted for widescale use in any program. SP remains the recommended drug regimen for IPTp, and countries are still reaching consensus on the regimen for IPTsc, with PDMC implementation currently being scaled up.^28^ However, WHO recommends reserving ACTs for clinical treatment of malaria due to concerns about emerging artemisinin resistance.^29^ SP and SP+AQ are also priced lower then DHA+PPQ: SP+AQ costing 0.43USD per treatment course versus 2.06USD for DHA+PPQ.^30^ Therefore the relative clinical impact of the drugs is essential to cost-effectiveness.^31–33^

As countries consider scaling up SMC and PMC in new locations with different resistance profiles, it is essential to leverage all existing evidence to guide decisions on which drug regimens are most suitable in these settings. Here, we conducted a systematic review and meta-analysis comparing the efficacy of SP+AQ for preventing malaria in children against other antimalarial drugs in different resistance settings, drawing on therapeutic efficacy and chemoprevention trials.

## Methods

### Search strategy and selection criteria

We performed a systematic review and meta-analysis in accordance with the Preferred Reporting Items for Systematic reviews and Meta-Analyses statement (PRISMA; appendix, p X), under PROSPERO registration CRD42024577149.^34^ We searched PubMed, Embase, MEDLINE, Global Health and the Worldwide Antimalarial Resistance Network (WWARN) Clinical Trials database^35^ for published randomised-controlled trials comparing SP+AQ with other antimalarials (Supplementary Table 1) from database inception until 06 December 2025 (search terms in Supplementary Methods, p 4).

Data were obtained from two main types of study: (1) chemoprevention studies, which provide preventive antimalarials to individuals regardless of their malaria infection status, then follow up individuals to assess incidence of malaria; and (2) therapeutic efficacy studies whose primary purpose is to assess adequate clinical and parasitological response to different antimalarials, but which also assess reinfection after treatment. For all studies and trial arms, we used the time at risk of new infection and the number of new infections or new clinical episodes to estimate the incidence rate ratio of SP+AQ against the comparator arms. Studies were included if they had a minimum of 28-day follow-up, and measured incidence after treatment of either clinical malaria episodes or new infections based on PCR-correction (full inclusion and exclusion criteria Supplementary Methods, p 4). In Covidence software, two independent reviewers (from GC-D, AM and OS) screened each study.^36^ Disagreements were resolved by consensus between reviewers.

### Data extraction and quality assessment

For a randomly selected 20% of full-text articles, extraction was done independently by the two reviewers (GC-D and AM). A consensus on discordant results was established before extraction of the remaining full texts. We extracted aggregated data from each of the included studies on the population, interventions arms and malaria outcomes (full list of variables appendix p A). All available case definitions, including severe malaria, all-cause hospital admissions and deaths were extracted (Supplementary Table 2). Two investigators (GC-D and OS) assessed the quality of studies according to an adapted version of the Cochrane Risk of Bias assessments for individual and cluster randomised controlled trials and stepped-wedge RCTs^37^ (full consolidated questionnaire Appendix pp 7 - 8), using a subset of 10% to test concordance of the two reviewers.

### Definition of drug resistance categories

For each included trial, data on key mutations associated with sulfadoxine, pyrimethamine and amodiaquine resistance were extracted^8,14,15,38^: *dhfr* N51I, C59R, S108N, I164L, *dhps* A437G, K540E, A581G, A613S/T and *mdr1* N86Y. We preferentially used mutation data from the trial itself if reported; in its absence, we used published systematic reviews of resistance markers^39,40^ and literature searches to identify mutation data close to the trial location and time (see Appendix pp 8 – 9),prioritising surveys within five years of the trial and 500km, with *n* ≥ 20 samples of *P. falciparum* parasites. Surveys satisfying these criteria were available for 95% (42/44) of study sites and years, but for the remaining 5% (2/44) of study sites and years (2/44) these boundaries had to be extended (Supplementary Table 4 and 5). For the trial in Madagascar, only molecular surveillance studies in country were considered due to its geographic isolation from mainland Africa. In some cases, *dhps* A581G was not surveyed in the drug resistance survey. In these instances, we use predicted prevalence from drug resistance mapping estimates^12^ to provide the prevalence of this marker. In all cases where use of predicted prevalence was necessary, it was 0%.

We categorised studies into settings of low, moderate and high SP resistance based on *dhps* A437G, K540E and A581G mutation prevalence (Supplementary Table 6). Thresholds were chosen based on the clustering of prevalences (Supplementary Figure 1 and 2). SP resistance was categorised as low if *dhps* K540E prevalence was < 30%, moderate if *dhps* K540E prevalence ≥ 30% and A581G prevalence < 5%, with high SP resistance classified as K540E prevalence ≥ 30% and A581G prevalence ≥ 5%. Amodiaquine resistance was categorised as low resistance where *mdr1* N86Y prevalence was < 40% based on previous studies and categorised as high elsewhere.^38^ We combined SP and AQ resistance, respectively, into six categories (Supplementary Table 3).

### Statistical analysis

The primary outcome of interest was the malaria incidence rate ratio (IRRs) between drug arms. Where available, we extracted the person-time-at-risk directly from the publication; where not explicitly provided, we inferred this based on the reported number of individuals at risk and censoring over time (Appendix pp 9 - 10). In therapeutic efficacy studies, we included only reinfections as assessed by PCR in the incidence metric and excluded recrudescent infections. In chemoprevention studies, the primary outcome was clinical episodes. We assumed for the meta-analysis that the percentage reduction in malaria incidence provided by each drug or drug combination would be equal whether measuring symptomatic episodes or all new malaria infections. This assumption is supported by available studies that measured both types of outcomes. For studies with multiple outcomes, we use the most stringent case definition based on a higher parasite density threshold where available and presence of symptoms. Due to the short half-life of artesunate, we assume that it does not contribute to the protective efficacy when combined with other antimalarials.^41^ Therefore, for the limited number of trials that have two arms that are treated as the same comparator (i.e. SP alone and SP+AS, AQ alone and AS+AQ), the arms were pooled together.

Meta-analyses and visualisations were undertaken using R package *meta*^42^ and all analyses were conducted in R (version 4.3.3).^43^ In all cases, common effects meta-analyses were undertaken, using the inverse-variance weighting on log-transformed IRRs and their corresponding standard errors (SEs). For the meta-analyses with ten or more data points, we additionally conducted a random-effects meta-analysis. Heterogeneity was measured by *I*^2^, with between-study variance, *τ*^2^, calculated using the DerSimonian-Laird estimator, with p-values for heterogeneity calculated according to the Q-test using Cochran’s Q-statistic. For sub-group analyses, sub-group specific pooled estimates are calculated for common (and random effects where appropriate), with sub-group differences assessed by the Q-statistic. Sub-group analyses were undertaken based on categories of SP and AQ resistance (Supplementary Table 3), case definitions (Supplementary Table 2), and the type of control arm (placebo or no placebo).

## Results

The search strategy returned 1,164 studies, from which we removed 435 duplicates (Figure 1). After abstract and full-text screening, 29 studies met the inclusion criteria: 12 chemoprevention effectiveness studies and 17 therapeutic efficacy studies (Table 1). The studies represented 13 countries across 32 unique sites and included 38,252 individual participants with 15,175 receiving SP+AQ and the remainder in one of 11 eligible comparator arms (Supplementary Table 1) (Figure 2).^44^ Some sites had studies in more than one year, giving a total of 44 studies for analysis. 26 studies had low SP resistance, 13 moderate SP resistance and the remaining 5 were classified as high SP resistant. A total of 23 studies were classified as having low AQ resistance and the remaining 21 studies as high AQ resistance (Supplementary Table 6).

**Figure 1:**
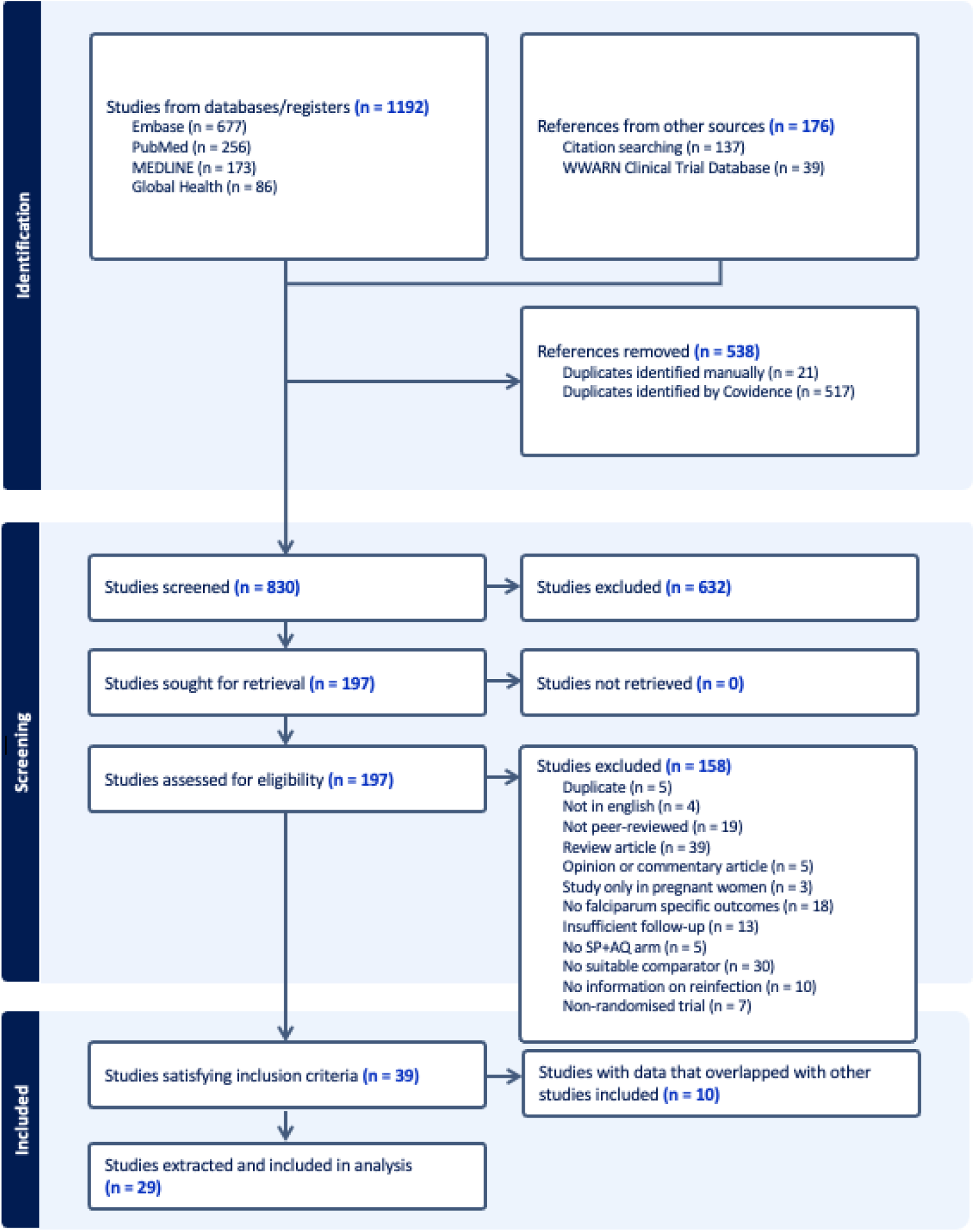
PRISMA Flowchart Flow diagram of study selection process. Full details of the search terms, inclusion and exclusion criteria are given in the Supplementary Methods.

**Figure 2.**
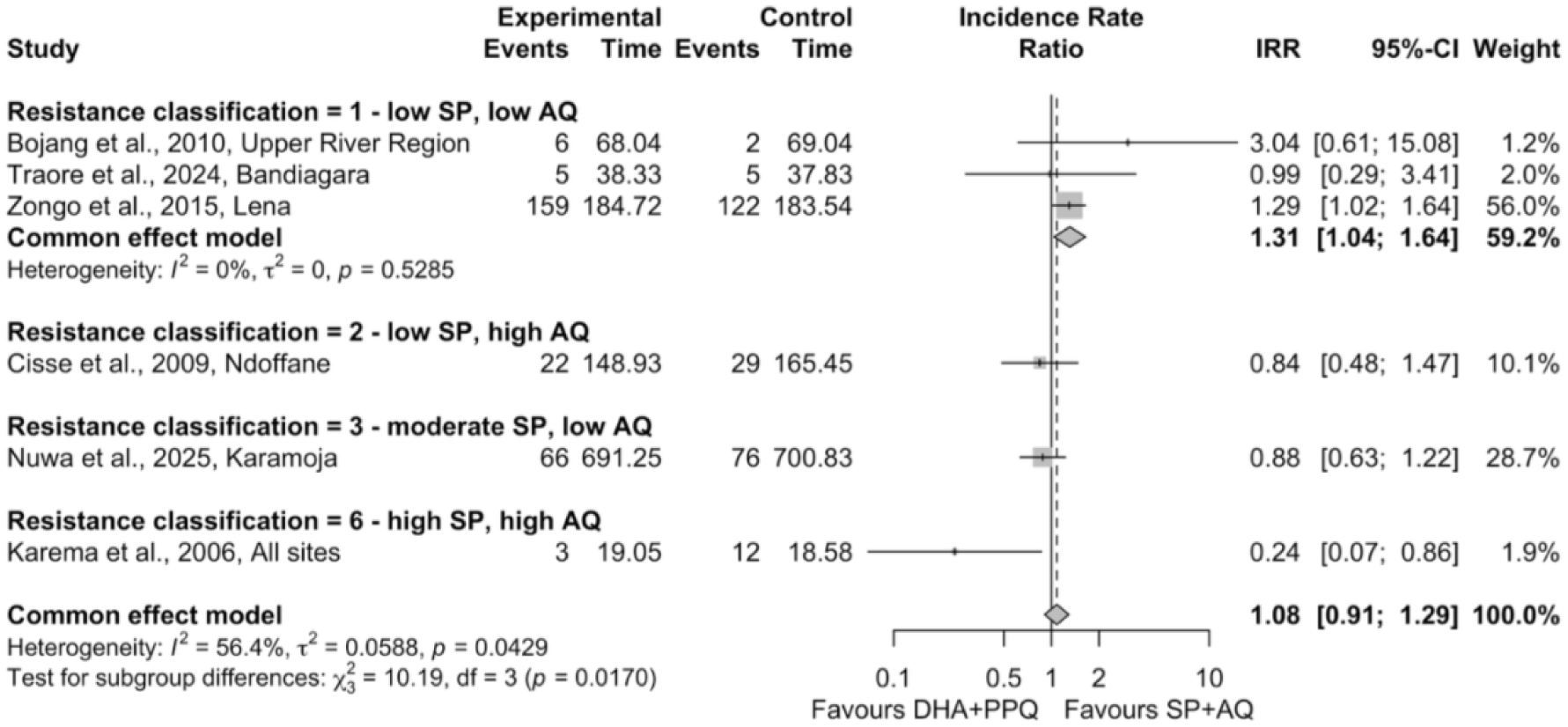
Common-effects meta-analysis of dihydroartemisinin-piperaquine (DHA+PPQ) versus sulfadoxine- pyrimethamine-amodiaquine (SP+AQ), stratified by drug resistance setting. Definitions of resistance classifications are found in Supplementary Table 3. SP resistance: low ∼ dhps K540E < 30%; moderate ∼ dhps K540E ≥30%, dhps A581G < 5%; high ∼ dhps K540E ≥30%, dhps A581G ≥ 5%; AQ resistance: low ∼ mdr1 N86Y < 40%; high ∼ mdr1 N86Y ≥ 40%.

**Table 1.**
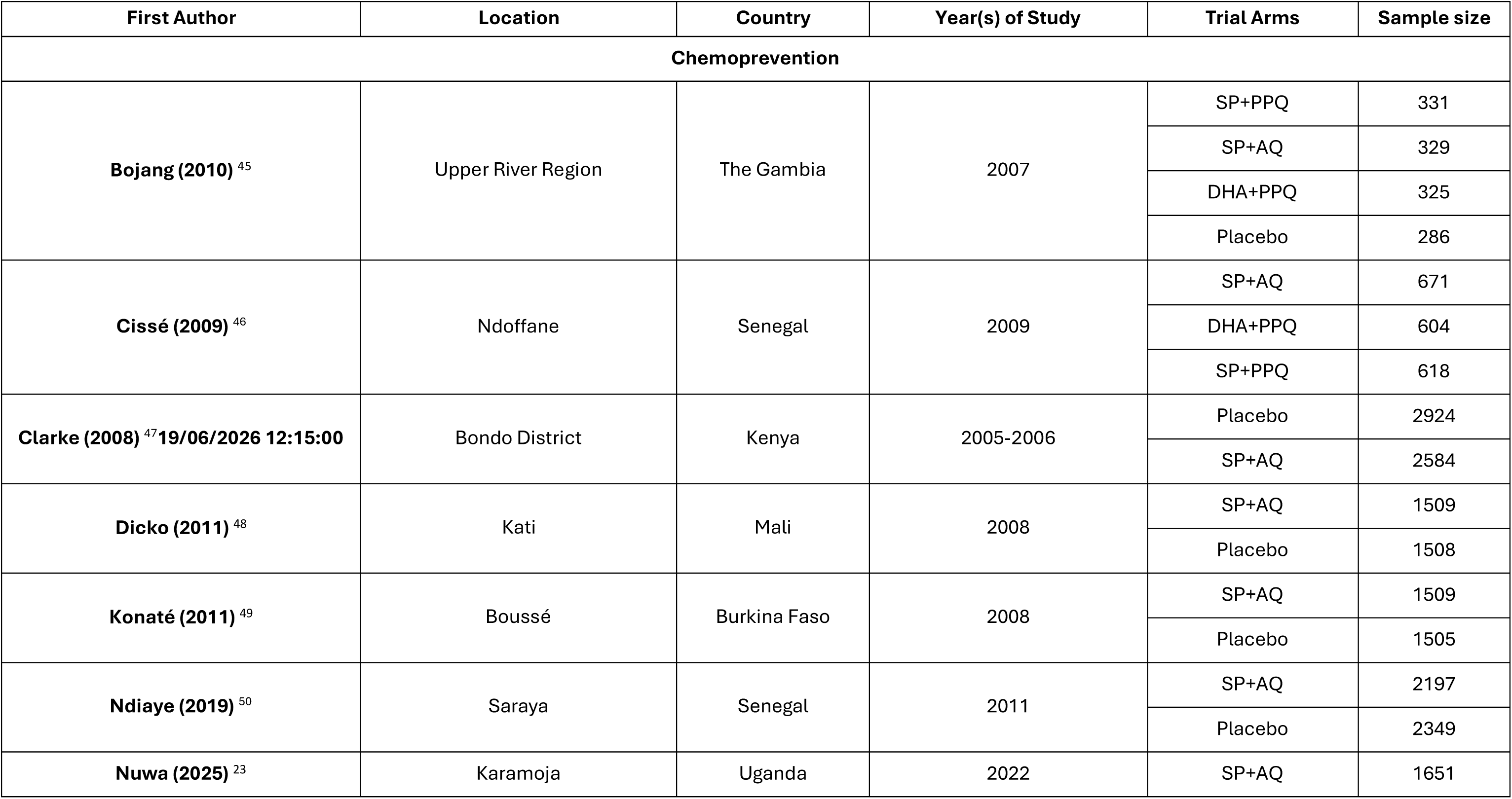
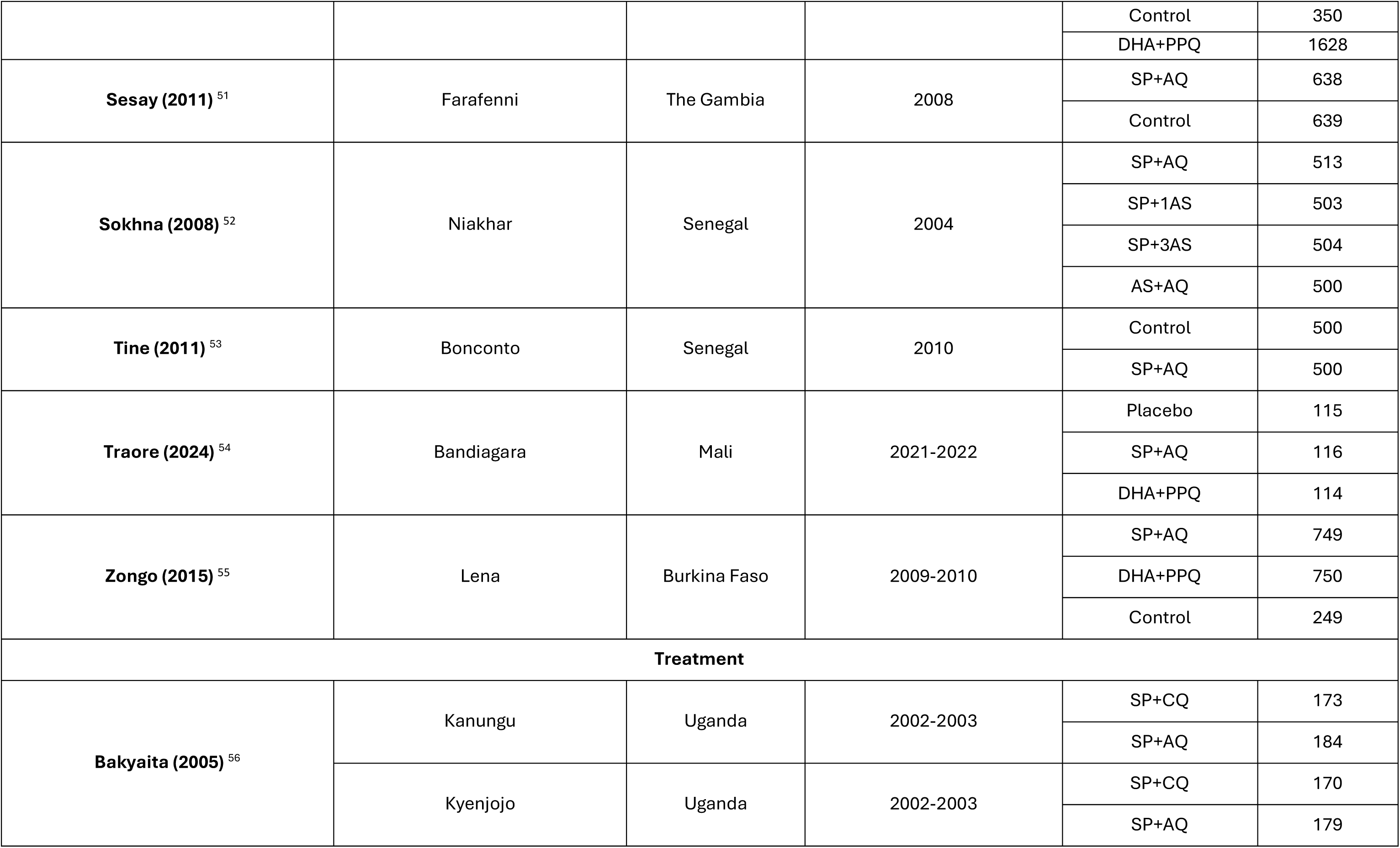
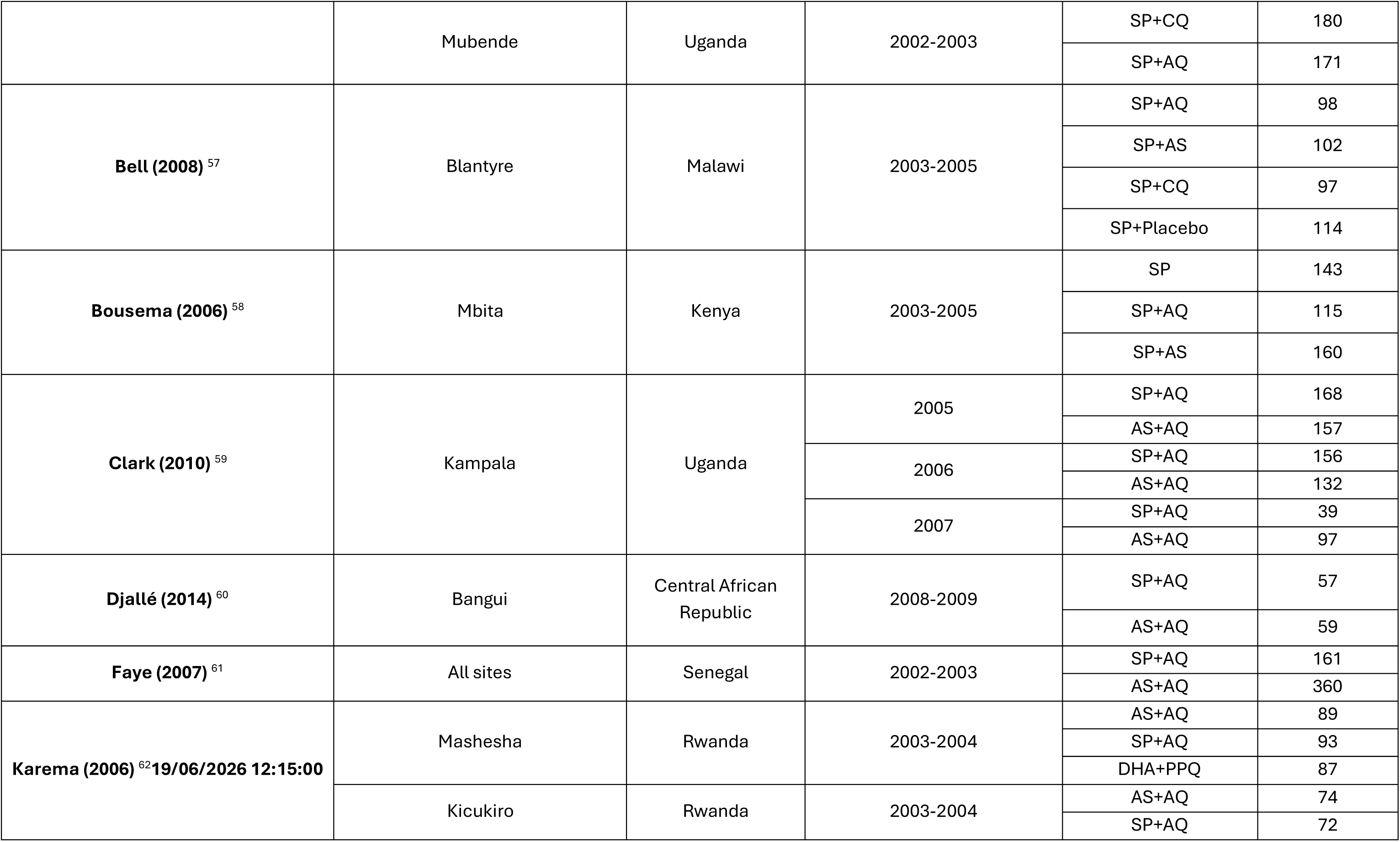
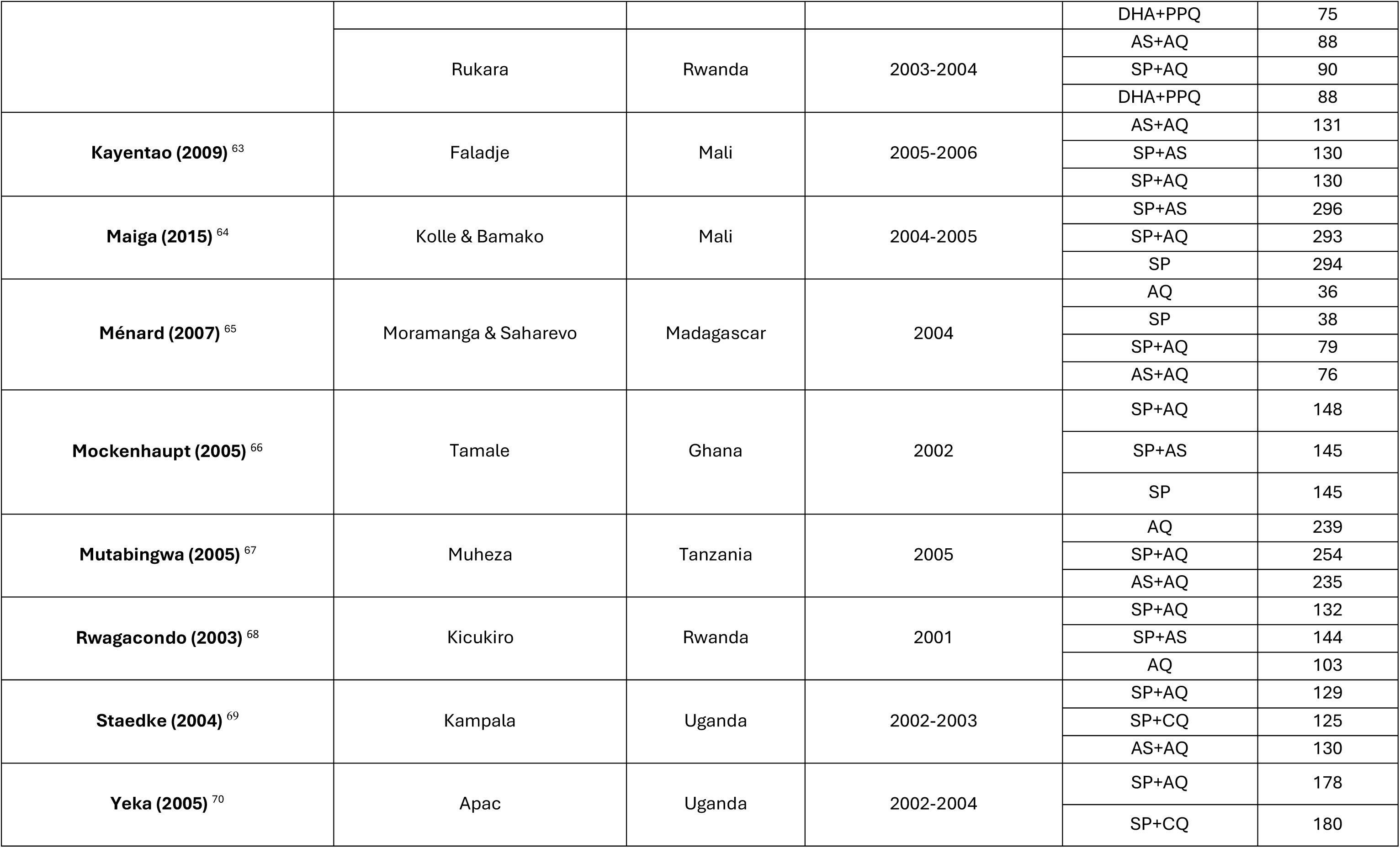
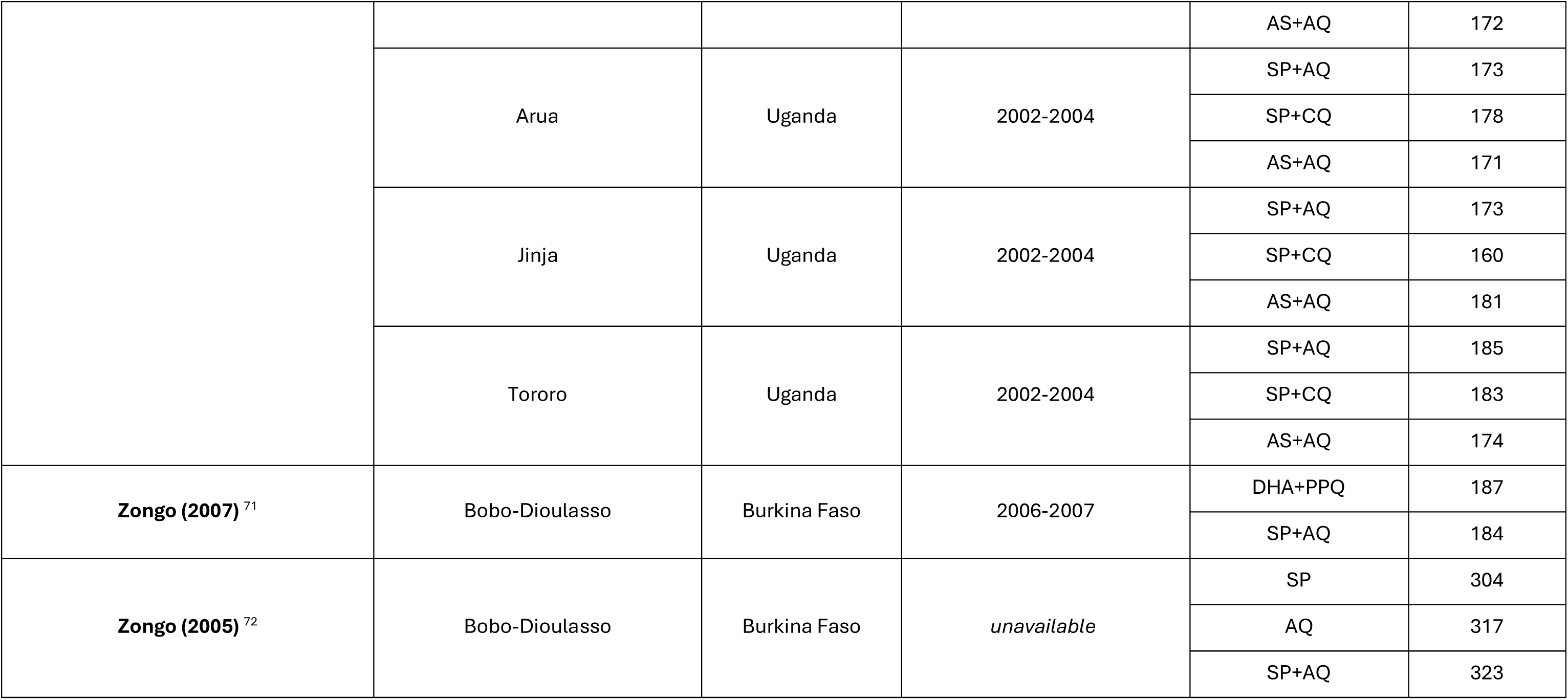
Studies included in the systematic review and meta-analysis. The chemoprevention section describes the 12 studies assessing malaria chemoprevention, which includes seasonal malaria chemoprevention in standard and extended age ranges. The treatment section describes the 17 therapeutic efficacy studies that satisfied the inclusion criteria. For multi-site trials, the sample size is split into each respective location. Similarly, for multi-year trials the arms and their sample size is shown for each year separately. The level of disaggregation in this table is in line with that throughout the meta-analysis both spatially and temporally.

Six studies compared SP+AQ against DHA+PPQ, two of the most commonly considered antimalarial combinations for chemoprevention. We estimated a pooled IRR of 1.08 (95% CI: 0.91, 1.29) comparing SP+AQ to DHA+PPQ in meta-analysis, indicating that DHA+PPQ provided comparable protection to SP+AQ, with a high degree of heterogeneity in estimates (*I^2^*=56.4)(Supplementary Figure 3). Subgroup analysis by SP and AQ resistance categories illustrates a trend that as resistance to SP and AQ increases, SP+AQ performs less well compared to DHA+PPQ although the number of studies in some resistance categories was small (Figure 2). In settings with low SP resistance (*dhps* K540E < 30%) and low AQ resistance (*mdr1* N86Y < 40%), we estimate that SP+AQ is superior to DHA+PPQ, averting 23.6% (95% CI: 3.8, 39.3%) of cases expected to occur after chemoprevention with DHA+PPQ. In settings with low-moderate combined resistance to SP and AQ, this combination is non-inferior to DHA+PPQ (Figure 2). In the one study where SP and AQ resistance are both high (*dhps* A581G prevalence > 5% and *mdr1* N86Y prevalence of > 40%), this showed that DHA+PPQ averted 75.6% (95% CI: 13.6, 93.1%) of cases expected to occur after chemoprevention with SP+AQ.

Seven studies compared either SP or SP+AS against SP+AQ. We assumed that artesunate does not contribute to the prophylactic protection of SP due to its very short half-life^41^ and therefore combined these arms in this meta-analysis. Overall, we estimate an IRR of 2.20 (95% CI: 1.51, 3.20), corresponding to SP+AQ averting an additional 54.6% (95% CI: 33.8, 68.8%) of cases compared to SP/SP+AS ( Figure 3). A subgroup meta-analysis showed that SP+AQ is more effective than SP/SP+AS in all resistance settings, including high SP and high AQ resistance settings ( Figure 3) although there were limited number of studies in moderate/high SP resistance settings.

**Figure 3.**
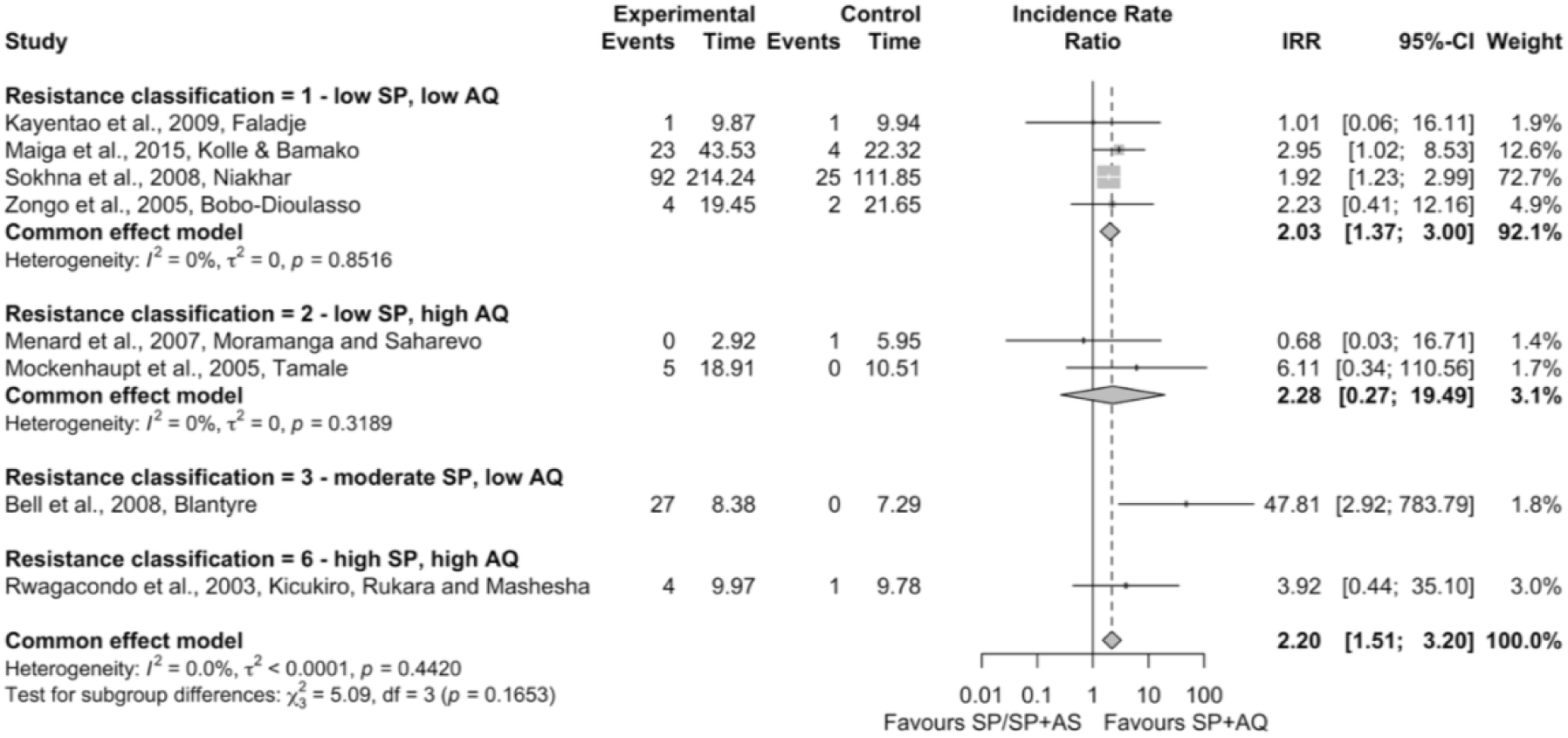
Common effects meta-analysis of sulfadoxine-pyrimethamine (SP) or sulfadoxine-pyrimethamine plus artesunate (SP+AS) versus sulfadoxine-pyrimethamine plus amodiaquine (SP+AQ), split by resistance setting |. Low, moderate and high indicate the levels of resistance to each drug. Full definitions of resistance classifications are found in Supplementary Table 3. SP resistance: low ∼ dhps K540E < 30%; moderate ∼ dhps K540E ≥30%, dhps A581G< 5%; high ∼ dhps K540E ≥30%, dhps A581G ≥ 5%; AQ resistance: low ∼ mdr1 N86Y < 40%; high ∼ mdr1 N86Y ≥ 40%. SP+1AS and SP+3AS are used to distinguish trial arms SP + one dose of AS per cycle (SP+1AS), versus 3 daily doses of AS per cycle (SP+3AS), but are treated as one comparator drug.** = data points that required a continuity correction

10 studies reported SP+AQ against either AQ alone or combined with artesunate (AS+AQ). SP+AQ was superior to AQ/AS+AQ in a common effects meta-analysis (Figure 4; IRR=1.26, 95% CI=1.10-1.43), and random effects meta-analysis (Supplementary Figure 10; IRR=1.36, 95% CI=1.01-1.81). We find statistically significant subgroup differences by SP resistance, with SP+AQ being superior to AQ/AS+AQ at low and moderate SP resistance (Figure 4). There were only 2 studies in settings with high SP resistance, however in both SP+AQ does not appear to offer additional protective efficacy against new infections (Figure 4; IRR=0.78, 95% CI: 0.34, 1.79).

**Figure 4.**
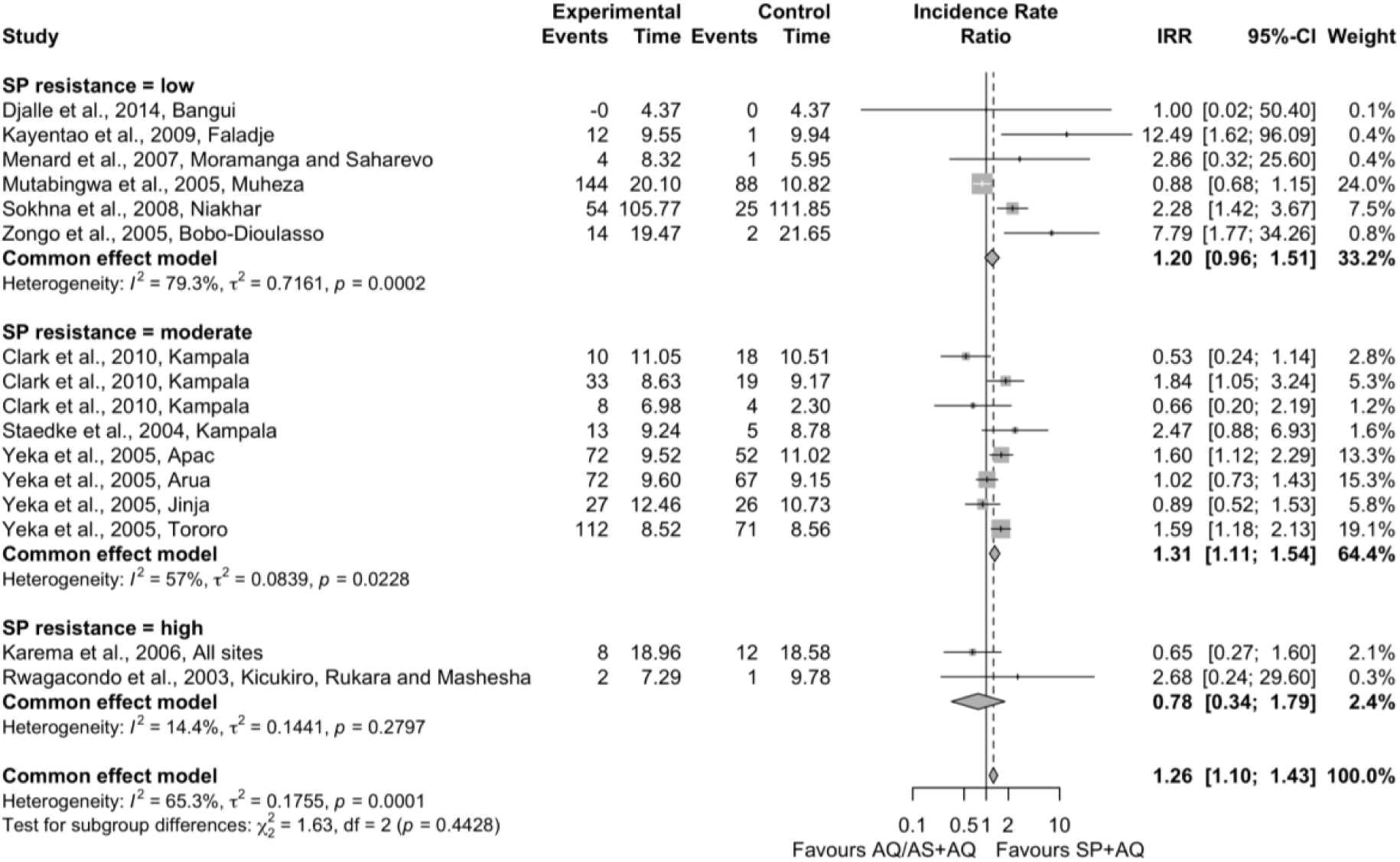
Common effects meta-analysis of amodiaquine (AQ) and artesunate-amodiaquine (AS+AQ) versus sulfadoxine-pyrimethamine plus amodiaquine (SP+AQ), split by SP resistance |. Definitions of resistance classifications are found in Supplementary Table 3. SP resistance: low ∼ dhps K540E < 30%; moderate ∼ dhps K540E ≥30%, dhps A581G < 5%; high ∼ dhps K540E ≥30%, dhps A581G ≥ 5%.

Four studies assessed SP+Chloroquine (SP+CQ) against SP+AQ across nine study sites. We estimate an IRR of 2.10 (95% CI: 1.82, 2.43), corresponding to SP+AQ averting an additional 52.3% (95% CI: 45.1, 58.8%) of cases compared to SP+CQ (Supplementary Figure 13).

For the eight studies that compared control versus SP+AQ, we estimated an IRR of 0.19 (95% CI: 0.18, 0.20) corresponding to SP+AQ averting 81.2% (95% CI: 79.9, 82.4%) of cases compared to the control (Supplementary Figure 17). A subgroup meta-analysis split by study design (placebo vs. non-placebo control), found a statistically significant subgroup difference between study designs, indicating that non-placebo-controlled trials had higher estimates of percentage of cases averted by SP+AQ (Supplementary Figure 18). A further meta-analysis by SP resistance classification showed a significant subgroup difference, but it is hard to distinguish if this is due to resistance alone or influenced by the differences in study design (Supplementary Figure 19). Additional meta-analyses estimated an IRR of 0.59 (95% CI: 0.33, 1.07) comparing SP+PPQ against SP+AQ, but this estimate was only based on two studies in Senegal and Gambia (Supplementary Figure 20).

## Discussion

Our meta-analysis finds SP+AQ to be a highly effective drug for chemoprevention across resistance settings. We find that SP+AQ was superior to SP alone (or in combination with AS which has negligible chemoprevention effect) for preventing new malaria infections and clinical cases in children, averting an estimated 54.5% of cases that would have occurred if SP was used on its own. This finding was true across different levels of resistance to SP or AQ, even in areas with low SP resistance. This result is surprising because previous estimates suggested the duration of protection provided by SP alone would be substantially longer than the duration of AQ protection when there is low SP resistance.^15,73,74^ The results presented here support the hypothesis that AQ plays an important role in SP+AQ prophylaxis, especially in areas with higher grade SP resistance.^57^ In these settings, the duration of protection offered by AQ may be similar or longer to that of SP alone, particularly when AQ resistance is low.^15^ There were, however, few studies in areas with higher levels of AQ resistance. Currently, molecular markers associated with AQ resistance have declined across nearly all malaria-endemic areas of Africa meaning that the current prevalence of *mdr1* N86Y is lower than at the time most of the studies were conducted.^40,75^ This means that we might expect SP+AQ to be more effective due to increased amodiaquine susceptibility, but in areas with high SP resistance may mean that AQ is the pharmacological agent providing the majority of the protection, raising concerns about parasite drug exposure.^76^

SP+AQ is already used for SMC, and our findings suggest strong consideration should be given to using SP+AQ for PMC in infants and young children rather than SP alone. A paediatric dispersible formulation of SP+AQ is available,^77^ however, the use of SP+AQ in a country will depend on other factors, in particular whether AS+AQ is being used for first-line case management, which may risk additional selection pressure for resistance against AQ. Historically, many countries implementing SMC changed first line treatment from AS+AQ to artemether-lumefantrine in order to avoid this problem, but the change may not be possible everywhere. These trade-offs must be carefully considered, since on the other hand, the reduction in burden associated with chemoprevention could reduce the number of curative malaria treatments required and hence potentially the propagation of resistance to artemisinin and/or partner drugs. AQ must also be given as 3 doses over 3 days, which leads to adherence challenges compared to SP alone which is a single dose. There are also issues of amodiaquine toxicity in breakthrough malaria cases.^71^

On average, SP+AQ also performed better than AQ alone (or AS+AQ), with a pooled estimate of 21% reduction in incidence, although in areas with high SP resistance there was no significant advantage, although the number of studies was small. Considering studies which compared SP+AQ to control groups, we found a similar effect size to a previous meta-analysis on this topic.^3^0 It was unclear whether variation in impact across areas was a result of drug resistance or study design because all studies in areas with higher resistance were unblinded, and on average the unblinded studies had a higher effect estimate than the blinded studies.

Our analysis found that SP+AQ was superior to DHA+PPQ in areas with low SP and low AQ resistance. However, in areas with moderate-to-high drug resistance to SP and/or AQ, there was a trend for DHA+PPQ to be non-inferior, and in the one study in an area of high SP and AQ resistance, DHA+PPQ was superior, although there were insufficient numbers of studies in these resistance settings to draw robust conclusions. Choice of an ACT for chemoprevention may require careful consideration due to emerging artemisinin resistance in Eastern and Southern Africa.^78–81^ WHO guidance in the response to antimalarial resistance in Africa indicates that chemoprevention and treatment drugs should use distinct antimalarial agents.^29^ Using SP+AQ for the chosen SMC drug regimen has several benefits. Firstly, it is a less costly drug than the main alternative DHA+PPQ ^30,31^. It additionally reserves the use of ACTs, such as DHA+PPQ as frontline treatment.

We reviewed studies in children, and our results are not intended to be applicable to intermittent preventive treatment against malaria in pregnancy (IPTp). Evidence from IPTp trials suggests that SP alone is still effective in preventing low birth weight and other adverse outcomes even in areas with SP resistance.^8,82^ The mechanisms by which pregnant women and their infants benefit from SP is considered to be very different, with a likely stronger role of non-malarial effects of SP in pregnant women.^83^ DHA+PPQ was found to be superior to SP in preventing malaria infection in pregnancy in recent trials in areas where there was high parasite resistance to SP, but it was inferior in preventing low birthweight specifically and adverse pregnancy outcomes more generally.^84^

Our review and meta-analyses drew data from treatment efficacy studies which measured reinfection as well as chemoprevention studies. This is a strength in increasing the evidence base for the effectiveness of different antimalarials, and in enabling use of data spanning a range of different levels of resistance to SP and AQ, particularly in areas with higher levels of *dhfr* and *dhps* mutation prevalence. However, there are some limitations to this approach. We assumed that protective efficacy of a drug as measured by percentage of reinfections prevented in treatment efficacy trials over 4-6 weeks would be approximately equivalent to the percentage of clinical episodes prevented by that drug in chemoprevention trials. This may be reasonable given that clinical episodes are associated with new infections,^85^ but our assumption would benefit from wider testing in future trials measuring both outcomes. In all treatment trials, infections occurring early during follow up (0 to 15 days) were assumed to be recrudescences and not included in this analysis. In addition, there were differences in the resistance levels by trial type with most chemoprevention studies undertaken in sites with low frequency of SP resistance-conferring mutations, and most treatment trials in areas of established SP drug resistance.

All except one included study was undertaken before 2013, which allowed inclusion of a broader range of SP and AQ resistance settings but also has limitations. For example, drug formulations for children have changed, with the dispersible formulation of SP+AQ introduced in 2012. While key resistance markers were captured in our review, more recently emerging variants such as the *dhps* 431V mutation, now common in Nigeria and Cameroon, could not be studied.^86^ Additional unknown genetic determinants of resistance, which are complex and likely not fully elucidated, may also have changed over time. Chemoprevention studies that were undertaken prior to the guidance for chemoprevention efficacy studies and therefore do not follow a standardised design like TES, which were standardised earlier.

Overall, our systematic review found evidence that SP+AQ performs well as paediatric malaria chemoprevention when compared against alternative drug regimens, across a variety of settings with different levels of resistance to each component drug. These findings support the retained usage of SP+AQ as a widely used malaria chemoprevention regimen in children, especially in the face of artemisinin resistance in Africa and the recommendations of WHO to reserve usage of ACTs for malaria treatment.^29^ Future work should prioritise comparison of the most used malaria chemoprevention drugs across resistance settings to provide more empirical evidence for phenotype-genotype associations for malaria chemoprevention.

## Data Availability

All data in the study are available via the GitHub repository.

https://github.com/ginacuomo/spaq_review/tree/main/data

