## Supplementary Information for "Comparative effectiveness of sulfadoxine–pyrimethamine plus amodiaquine versus other antimalarial regimens for paediatric malaria chemoprevention in the context of drug resistance: a systematic review and meta-analysis"

### Table of Supplementary Figures

Supplementary Figure 1 | Figure illustrating the prevalence of *dhps* K540E and A581G across the study sites. Dashed lines indicate the thresholds defining SP resistance categories (low/moderate/high). ........ 14

Supplementary Figure 2 | Histogram of *mdr1* N86Y prevalence. Dotted vertical line represents the cut off between low and high AQ resistance. ........ 14

Supplementary Figure 3 | Meta-analysis comparing DHA+PPQ vs SP+AQ. ........ 28

Supplementary Figure 4 | Meta-analysis comparing DHA+PPQ vs SP+AQ, split by SP resistance classifications as described in Supplementary Table 3 ........ 28

Supplementary Figure 5 | Meta-analysis comparing DHA+PPQ vs SP+AQ, split by AQ resistance classifications as described in Supplementary Table 3 ........ 29

Supplementary Figure 6 | Meta-analysis comparing DHA+PPQ vs SP+AQ, with subgroup analysis split by the study design. ........ 29

Supplementary Figure 7 | Meta-analysis of SP+AQ versus SP / SP+AS. ........ 29

Supplementary Figure 8 | Meta-analysis comparing SP / SP+AS versus SP+AQ, split by SP resistance classifications as described in Supplementary Table 3 ........ 29

Supplementary Figure 9 | Meta-analysis comparing SP / SP+AS versus SP+AQ, split by AQ resistance classifications as described in Supplementary Table 3 ........ 30

Supplementary Figure 10 | Meta-analysis comparing AQ / AS+AQ versus SP+AQ ........ 30

Supplementary Figure 11 | Meta-analysis comparing AQ / AS+AQ versus SP+AQ, with subgroup analysis by the resistance classification thresholds described in Supplementary Table 3 ........ 31

Supplementary Figure 12 | Meta-analysis comparing AQ / AS+AQ versus SP+AQ, with subgroup analysis by the SP resistance thresholds described in Supplementary Table 3. ........ 31

Supplementary Figure 13 | Meta-analysis comparing SP+CQ versus SP+AQ. ........ 32

Supplementary Figure 14 | Meta-analysis comparing SP+CQ versus SP+AQ, split by SP resistance as described in Supplementary Table 3. ........ 32

Supplementary Figure 15 | Meta-analysis comparing SP+CQ versus SP+AQ, split by AQ resistance as described in Supplementary Table 3. ........ 33

Supplementary Figure 16 | Meta-analysis comparing SP+CQ versus SP+AQ, split by resistance classification thresholds as described in Supplementary Table 3. ............ 33
Supplementary Figure 17 | Meta-analysis of SP+AQ versus control arms. .................. 34
Supplementary Figure 18 | Meta-analysis of SP+AQ versus control arms, split by trial design. ........................................................................................................ 34
Supplementary Figure 19 | Meta-analysis of SP+AQ versus control arms, split by resistance classification as described in Supplementary Table 3. ............................. 35
Supplementary Figure 20 | Meta-analysis of SP+PPQ versus SP+AQ.......................... 35

### Table of Supplementary Tables

Supplementary Table 1: Table illustrating eligible control arms for inclusion in the systematic review | Arm represents the possible descriptions of the trial arm in the studies themselves. Comparator lists the eligible comparator arms that trial arms map onto. *For the Sokhna study, there were two SP+AS arms with one and three doses of AS per cycle, respectively. ........ 6

Supplementary Table 2 | Table illustrating how the malaria outcomes reported in the included studies were mapped to a consistent and unified set of malaria outcomes. ... 9

Supplementary Table 3 | Classification of resistance categories ........ 13

Supplementary Table 4 | For each study site and year, a summary of the resistance marker information for SP mutations. For each study site and year, the geographic and temporal distance of the chosen marker survey are chosen, with time difference given in absolute years and distance calculated using the respective longitude and latitude of the trial site and marker survey. The PubMed ID of the chosen surveys are given, along with the prevalence of the three key mutations in the *dhps* gene. ........ 19

Supplementary Table 5 | For each study site and year, a summary of the resistance marker information for AQ mutations. For each study site and year, the geographic distance of the chosen marker survey are chosen, with time difference given in absolute years and distance calculated using the respective longitude and latitude of the trial site and marker survey. The PubMed ID of the chosen surveys are given, along with the prevalence of the *mdr1* N86Y mutation. ........ 22

Supplementary Table 6 | A table showing all study sites and years and the respective classifications with respect to SP, AQ and combined resistance classifications as described in Supplementary Table 3. ........ 27

### Supplementary Methods:

#### Search terms

We searched on Embase + Embase Classic, Medline and Global Health databases in the OVID platform using the following search terms:

(sulfadoxine.mp. or Sulfadoxine/ or sulphadoxine.mp. or pyrimethamine.mp. or Pyrimethamine/ or fansidar.mp) and (amodiaquine.mp. or Amodiaquine/) and (malaria.mp. or Malaria/ or Malaria, Falciparum/) and (clinical trial.mp. or Clinical Trial/ or longitudinal study.mp. or Longitudinal Studies/)

We searched PubMed using the following:

((((clinical trial*) OR (longitudinal stud*)) AND ((malaria) OR (plasmodium falciparum))) AND (amodiaquine)) AND ((((sulfadoxine) OR (sulphadoxine)) OR (pyrimethamine)) OR (fansidar))

#### Inclusion and exclusion criteria

At the title and abstract screening stage, the inclusion criteria was for studies that assessed *Plasmodium falciparum* malaria, assessed malaria in children, was undertaken in Africa, and received SP+AQ. If any of these criteria were ambiguous, we retained them for full-text review. We also did not assess if there was a suitable comparator arm at this initial stage.

The exclusion criteria for full-text review were ranked to reduce the number of conflicts in the full-text review stage, with the following exclusion criteria.

1. Duplicate record
2. Not in English
3. Not peer-reviewed
4. Review article
5. Opinion or commentary article
6. Study conducted outside Africa
7. Study only in pregnant women

8. No *falciparum*-specific outcomes
9. Not sufficient follow-up (<28 days)
10. No SP+AQ arm
11. No suitable comparator (see Table X)
12. No information on reinfection (see section X)

Suitable comparators are those that were one of the following: control, SP, SP+Placebo, SP+AS, SP+CQ, SP+PPQ, AS+AQ or DHA+PPQ. All trials were required to have an SP+AQ arm. This is expanded on in the table below, demonstrating the generic arms (which were matched against the list above), and what comparator arms would satisfy this (Table X).

| Arm | Comparator |
|---|---|
| Non-placebo control | control |
| Albendazol control | control |
| Placebo control | control |
| SP+AQ | SP+AQ |
| SP | SP |
| SP+Placebo | SP+Placebo |
| SP+AS | SP+AS |
| SP+1AS* | SP+AS |
| SP+3AS* | SP+AS |
| SP+CQ | SP+CQ |
| SP+PPQ | SP+PPQ |
| DHA+PPQ | DHA+PPQ |
| AS+AQ | AS+AQ |

**Supplementary Table 1: Table illustrating eligible control arms for inclusion in the systematic review | Arm represents the possible descriptions of the trial arm in the studies themselves.** Comparator lists the eligible comparator arms that trial arms map onto. *For the Sokhna study, there were two SP+AS arms with one and three doses of AS per cycle, respectively.

When multiple studies published information from the same trial, the publication presenting the most complete information necessary for our analysis was retained. When studies published data with multiple levels of resolution, we prioritised data with the finest temporal and spatial resolution and where the clinical outcome of the trial itself was available.

#### Data extraction

For therapeutic efficacy studies, we additionally extracted information on: the period of follow-up for which PCR correction was performed, how reinfection or recrudescence was defined according to the PCR results, PCR-corrected and PCR-uncorrected therapeutic efficacy and the reference to the case definition, as the meaning of these terms changed with updated World Health Organization therapeutic efficacy study guidelines.  For chemoprevention studies, we extracted information on other malaria interventions co-delivered, whether there was a randomised comparatorarm, type of control, the number of malaria chemoprevention cycles and their spacing, the number of cases and the child years at risk in the trial.

Across both types of study design, there was a wide range of malaria outcomes for included studies. These malaria outcomes were mapped to consistent categories to simplify this and are fully described in Supplementary Table 2.

| Case Definition | Grouping | Trial Design |
|---|---|---|
| Incidence with any parasitaemia | Clinical incidence (any parasitaemia) | chemoprevention |
| Incidence with any parasitaemia >= 5000/ul | Clinical incidence (parasitaemia >= 5000/ul) | chemoprevention |
| Incidence with any parasitaemia ITT | Clinical incidence (any parasitaemia) | chemoprevention |
| Incidence with any parasitaemia PP | | chemoprevention |
| Incidence with parasitaemia (>3000/ul) ITT | Clinical incidence (parasitaemia >= 3000/ul) | chemoprevention |
| Incidence with parasitaemia (>3000/ul) PP | | chemoprevention |
| Prevalence of infection - ITT | Asymptomatic incidence (any parasitaemia) | chemoprevention |
| Prevalence of infection - PP | | chemoprevention |
| Fever and asexual parasitaemia | Clinical incidence (any parasitaemia) | chemoprevention |

| | | |
|---|---|---|
| Clinical malaria defined as fever and parasitaemia >= 5000/ul | Clinical incidence (parasitaemia >= 5000/ul) | chemoprevention |
| Severe malaria | Severe malaria | chemoprevention |
| Clinical malaria = fever and any parasitaemia | Clinical incidence (any parasitaemia) | chemoprevention |
| Severe malaria (WHO 2000) | Severe malaria | chemoprevention |
| All-cause hospital admission | All-cause hospital admission | chemoprevention |
| Deaths | Deaths | chemoprevention |
| Malaria case (RDT confirmed) | Clinical incidence (RDT positive) | chemoprevention |
| Deaths from all causes | Deaths | chemoprevention |
| Clinical malaria at outpatient department | Clinical incidence (any parasitaemia) | chemoprevention |
| Fever and parasitaemia >= 3000/ul | Clinical incidence (parasitaemia >= 3000/ul) | chemoprevention |
| Fever and RDT positive | Clinical incidence (RDT positive) | chemoprevention |
| Symptoms + asexual parasitaemia (thick smear) | Clinical incidence (any parasitaemia) | chemoprevention |
| Fever with any parasitaemia | Clinical incidence (any parasitaemia) | chemoprevention |
| RDT confirmed malaria | Clinical incidence (RDT positive) | chemoprevention |
| Clinical failure | Clinical incidence (any parasitaemia) | treatment |
| Parasitological failure | Asymptomatic incidence (any parasitaemia) | treatment |
| Reinfection | Asymptomatic incidence (any parasitaemia) | treatment |

| | | |
|---|---|---|
| Early treatment failure | Clinical incidence (any parasitaemia) | treatment |
| Late treatment failure | Clinical incidence (any parasitaemia) | treatment |
| Treatment failure - all outcomes combined | Asymptomatic incidence (any parasitaemia) | treatment |
| Late clinical failure | Clinical incidence (any parasitaemia) | treatment |
| Late parasitological failure | Asymptomatic incidence (any parasitaemia) | treatment |
| Total treatment failure | Asymptomatic incidence (any parasitaemia) | treatment |

**Supplementary Table 2 | Table illustrating how the malaria outcomes reported in the included studies were mapped to a consistent and unified set of malaria outcomes.**

#### Quality assessment

We used an adapted version of the Cochrane Risk of Bias tool for individual-, cluster-randomised trials and stepped-wedge cluster randomised trials. This quality assessment was designed to assess bias introduced by the randomisation process, deviation from intended interventions, missing outcome data, measurement of the outcome and selection of reported results. Studies were assessed in all domains as (1) low risk, (2) some concerns and (3) high risk with questions adapted to trial design. The questions are summarised below:

Domain 1: Bias due to the randomisation process

- Was the allocation sequence **randomly generated**? (e.g., computer-generated, random number table)
- Was allocation **concealed** until recruitment was complete? *(e.g., central randomization, sealed envelopes)*
- (Cluster RCTs only): Was the randomization conducted **before cluster recruitment**? (Pre-randomization prevents selection bias)

Domain 2: Bias due to deviations from intended interventions

- Were trial personnel (e.g., clinicians, researchers) blinded to intervention assignment? If blinding was not feasible, were there systematic differences in how groups were treated beyond the intervention itself?
- Were participants blinded to their intervention assignment? If blinding was not feasible, were deviations from the intended intervention—*such as differences in adherence, access to additional malaria treatments, or variations in co-medications*—balanced between groups?
- *(Cluster RCTs only):* Were clusters or steps aware of their intervention status, potentially leading to contamination? (referring to members of the control group receiving the intervention, rather than contamination within the level of randomisation i.e. household, village etc.)
- Were there any systematic differences in care, co-interventions, or other contextual factors between groups that could influence outcomes (e.g., differential access to healthcare, additional treatments, **health information** or provider behaviour)?

Domain 3: Bias due to missing outcome data

- Were missing outcome data balanced across intervention groups? *(Consider dropout rates and reasons for missing data)*
- *(Cluster RCTs only)* Were clusters lost to follow-up?

Domain 4: Bias in measurement of the outcome

- Were outcome assessors **blinded** to intervention assignment?
- Was a **clear, pre-specified definition** of each outcome used, and were the **same measurement methods**, instruments, and timing applied consistently across all groups?

Domain 5: Bias in selection of reported results

- Was the trial protocol available, and were all pre-specified outcomes reported?

#### Definition of resistance categories

For each study site and year, the nearest possible resistance survey was identified. We assessed the distribution of prevalence of key markers. We looked at the prevalence of *dhps* A437G, K540E and A581G to determine clusters of prevalence of these specific markers. The definitions of the resistance classification were chosen to fall between the clusters of prevalence levels. Likewise, for amodiaquine resistance, we chose a threshold that divided the *mdr1* N86Y mutations into two clear clusters.

#### Estimating time at risk

The primary outcome of interest was the malaria incidence rate ratio (IRR) between drug arms. For all studies and trial arms, we used the time at risk of new infection and the number of new infections to estimate the incidence rate ratio of SP+AQ against the comparator arms.

For chemoprevention effectiveness trials and TES trials, distinct methods are required for inferring the time at risk and number of infections. For chemoprevention effectiveness trials, the number of cases, new infections and corresponding time-at-risk of infection are usually reported explicitly in the paper, therefore they generally do not require inference. When time-at-risk is not reported explicitly in chemoprevention studies, we inferred this based on the number of individuals at risk over time.

TES studies all had to be PCR-corrected in order to satisfy our inclusion criteria. PCR-corrected failure rate represents recrudescence, whereas PCR-uncorrected rate captures the sum of recrudescence and reinfection. The endpoint of interest to assess how antimalarials prevent against new infections is reinfection, and therefore we calculate the number of new infections as:

$$n_{reinfected} = (PCR_{uncorrected} - PCR_{corrected}) \cdot n_{outcome}$$

where $PCR_{uncorrected}$ is the proportion of participants with treatment failure by day of follow-up (recrudescence + reinfection), $PCR_{corrected}$ is the proportion with confirmed recrudescence, and $n_{outcome}$ is the number of participants with an outcome recorded at the end of follow-up.

It was standard in TES trials to perform PCR correction for a specified period of follow-up. Before the start of PCR-correction, the trial assumes that any symptomatic or asymptomatic malaria infection is a result of recrudescence of day 0 parasitaemia. Therefore, individuals are only at risk of new infection, as defined by the trial, during the period in which PCR-correction is conducted. We assume that reinfection is equally likely throughout the duration of the trial and only consider individuals to be at risk of reinfection new infection for the dates where PCR-correction would be performed. Outside of these days of follow-up, reinfections would be incorrectly identified as recrudescent infections and therefore would not be captured in our calculation of the number of reinfections. Time at risk for TES studies is inferred as:

$$t_{risk} = \frac{PCR_{end} - PCR_{start}}{2} \cdot n_{reinfected} + t_{follow-up} \cdot n_{uninfected}$$

Where $t_{risk}$ is the inferred time at risk, $PCR_{start}$ and $PCR_{end}$ are the start and end of PCR-correction, respectively, $t_{follow-up}$ is the duration of follow-up, and $n_{uninfected}$ $and$ $n_{reinfected}$ are the number of study participants who remain uninfected at the end of follow-up or are reinfected, respectively.

### Supplementary Results

#### Resistance Markers and Classifications

The combined SP/AQ resistance classifications are simply all the possible combinations of the SP and AQ resistance categories respectively. Supplementary Figures 1 and 2 show how the prevalence of mutations clustered together and Supplementary Table 3 shows the resistance classifications. Supplementary Table 4 and 5 shows the prevalence of the mutations in each of the key markers for each trial site and year, and the corresponding resistance classifications are shown in Supplementary Table 6.

| Classification | *dhps* K540E | *dhps* A581G | mdr1 N86Y |
|---|---|---|---|
| Low SP resistance | <30% | 0% | NA |

| | | | |
|---|---|---|---|
| Moderate SP resistance | >30% | 0% | NA |
| High SP resistance | >30% | >5% | NA |
| Low AQ resistance | NA | NA | <40% |
| High AQ resistance | NA | NA | >40% |
| Low SP, low AQ resistance (class 1) | <30% | 0% | <40% |
| Low SP, high AQ resistance (class 2) | <30% | 0% | >40% |
| Moderate SP, low AQ resistance (class 3) | >30% | 0% | <40% |
| Moderate SP, high AQ resistance (class 4) | >30% | 0% | >40% |
| High SP, low AQ resistance (class 5) | >30% | 5% | <40% |
| High SP, high AQ resistance (class 6) | >30% | 5% | >40% |

**Supplementary Table 3 | Classification of resistance categories**

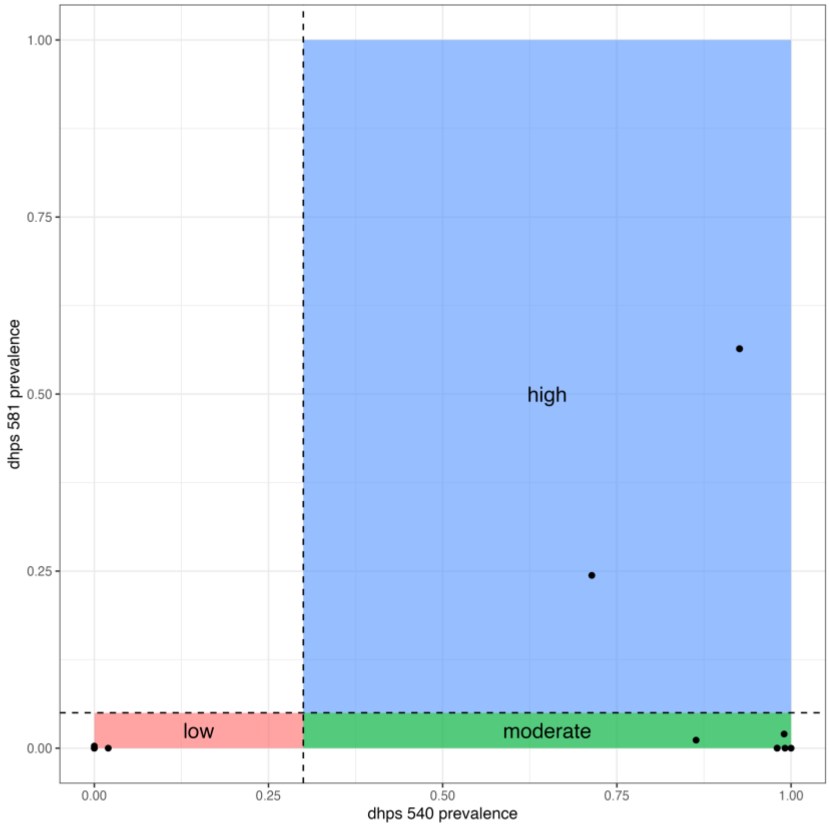

**Supplementary Figure 1 | Figure illustrating the prevalence of *dhps* K540E and A581G across the study sites.** Dashed lines indicate the thresholds defining SP resistance categories (low/moderate/high).

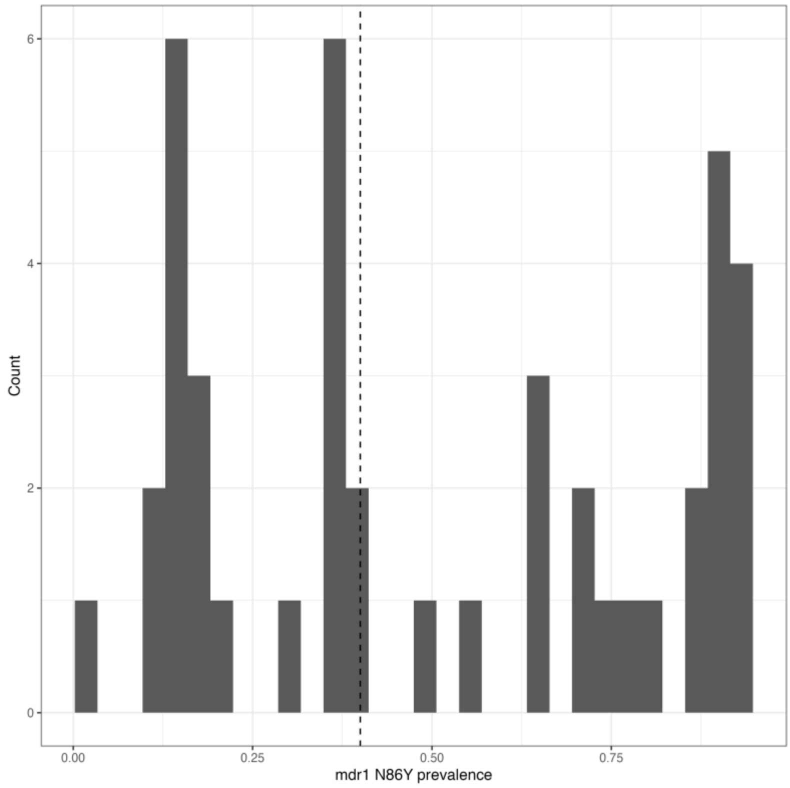

**Supplementary Figure 2 | Histogram of *mdr1* N86Y prevalence.** Dotted vertical line represents the cut off between low and high AQ resistance.

| Author | Year start | Year end | location | country | Distance (km) | Difference (years) | PMID | *dhps* 437 | *dhps* 540 | *dhps* 581 |
|---|---|---|---|---|---|---|---|---|---|---|
| **Bakyaita** | 2002 | 2003 | Kanungu | Uganda | 519.9 | 1 | 24799371 | 98.0% | 98.0% | 0.0% |
| **Bakyaita** | 2002 | 2003 | Kyenjojo | Uganda | 394.8 | 1 | 24799371 | 98.0% | 98.0% | 0.0% |
| **Bakyaita** | 2002 | 2003 | Mubende | Uganda | 311.8 | 1 | 24799371 | 98.0% | 98.0% | 0.0% |
| **Bell** | 2002 | 2005 | Blantyre | Malawi | 175.4 | 1 | 23198734 | 90.9% | 86.4% | 1.1% |
| **Bousema** | 2003 | 2004 | Mbita | Kenya | 34.3 | 2.5 | 22096496 | 92.5% | 82.3% | NA |
| **Clark** | 2005 | 2005 | Kampala | Uganda | 183.1 | 0 | 24799371 | 98.0% | 99.0% | 2.0% |
| **Clarke** | 2005 | 2006 | Bondo District | Kenya | 15.1 | 1.5 | 25889220 | 98.0% | 100.0% | 0.0% |
| **Karema** | 2003 | 2004 | All sites | Rwanda | 76 | 5 | 19841150 | 98.7% | 92.6% | 56.4% |

| | | | | | | | | | | |
|---|---|---|---|---|---|---|---|---|---|---|
| **Karema** | 2003 | 2004 | Kicukiro | Rwanda | 46.9 | 5 | 19841150 | 98.7% | 92.6% | 56.4% |
| **Karema** | 2003 | 2004 | Mashesha | Rwanda | 0.0 | 5 | 19841150 | 89.7% | 71.4% | 24.4% |
| **Karema** | 2003 | 2004 | Rukara | Rwanda | 0.0 | 5 | 19841150 | 98.7% | 92.6% | 56.4% |
| **Rwagacondo** | 2001 | 2001 | Kicukiro, Rukara and Mashesha | Rwanda | 76.0 | 5 | 19841150 | 98.7% | 92.6% | 56.4% |
| **Menard** | 2004 | 2004 | Moramanga and Saharevo | Madagascar | 146.2 | 2 | 19704124 | 36.1% | 0.0% | 0.3% |
| **Mutabingwa** | 2002 | 2004 | Muheza | Tanzania | 178.8 | 0.5 | 28240422 | 64.0% | 0.0% | 0.0% |
| **Nuwa** | 2022 | 2022 | Karamoja | Uganda | 0 | 0 | 39826559 | 100.0% | 99.1% | 0.0% |
| **Staedke** | 2002 | 2003 | Kampala | Uganda | 182.9 | 1 | 24799371 | 98.0% | 98.0% | 0.0% |
| **Yeka** | 2002 | 2004 | Apac | Uganda | 0 | 0 | 16518760 | 95.0% | 95.0% | NA |

| | | | | | | | | | | |
|---|---|---|---|---|---|---|---|---|---|---|
| **Yeka** | 2002 | 2004 | Arua | Uganda | 0 | 0 | 16518760 | 94.0% | 87.0% | NA |
| **Yeka** | 2002 | 2004 | Jinja | Uganda | 0 | 0 | 16518760 | 94.0% | 92.0% | NA |
| **Yeka** | 2002 | 2004 | Tororo | Uganda | 0 | 0 | 16518760 | 98.0% | 98.0% | NA |
| **Bojang** | 2007 | 2007 | Upper River Region | The Gambia | 69.9 | 3 | 23589534 | 44.6% | 0.0% | 0.0% |
| **Cisse** | 2007 | 2007 | Ndoffane | Senegal | 124.8 | 2 | 23617576 | 67.6% | 0.0% | NA |
| **Dicko** | 2008 | 2008 | Kati | Mali | 286.6 | 4 | 27662368 | 61.3% | 3.1% | NA |
| **Dicko** | 2008 | 2008 | Djoliba | Mali | 291.3 | 4 | 27662368 | 61.3% | 3.1% | NA |
| **Dicko** | 2008 | 2008 | Siby | Mali | 311.5 | 4 | 27662368 | 61.3% | 3.1% | NA |
| **Dicko** | 2008 | 2008 | Ouelessebougou | Mali | 269.9 | 4 | 27662368 | 61.3% | 3.1% | NA |

| | | | | | | | | | | |
|---|---|---|---|---|---|---|---|---|---|---|
| **Djalle** | 2008 | 2009 | Bangui | Central African Republic | 0 | 4.5 | 16474071 | 18.6% | 5.2% | |
| **Kayentao** | 2005 | 2006 | Faladje | Mali | 275.5 | 4.5 | 28381273 | 38.0% | 2.0% | 0.0% |
| **Konate** | 2008 | 2008 | Toeghin | Burkina Faso | 52.1 | 4 | 26368675 | 34.2% | 0.0% | NA |
| **Konate** | 2008 | 2008 | Niou | Burkina Faso | 29.3 | 4 | 26368675 | 34.2% | 0.0% | NA |
| **Konate** | 2008 | 2008 | Laye | Burkina Faso | 47. 7 | 4 | 26368675 | 34.2% | 0.0% | NA |
| **Konate** | 2008 | 2008 | Sao | Burkina Faso | 137.3 | 4 | 26368675 | 34.2% | 0.0% | NA |
| **Konate** | 2008 | 2008 | Bousse | Burkina Faso | 33.0 | 4 | 26368675 | 34.2% | 0.0% | NA |
| **Maiga** | 2004 | 2005 | Kolle & Bamako | Mali | 344.5 | 5 | 26486699 | 55.4% | 0.0% | 0.0% |
| **Mockenhaupt** | 2002 | 2002 | Tamale | Ghana | 90.7 | 3.5 | 23045251 | 80.0% | 0.0% | NA |

| | | | | | | | | | | |
|---|---|---|---|---|---|---|---|---|---|---|
| **Ndiaye** | 2011 | 2011 | Saraya | Senegal | 54.8 | 0 | 30865632 | 39.1% | 0.0% | 0.0% |
| **Sesay** | 2008 | 2008 | Farafenni | The Gambia | 126.2 | 1 | 23589534 | 44.6% | 0.0% | 0.0% |
| **Sokhna** | 2006 | 2006 | Niakhar | Senegal | 0.0 | 0 | 18213379 | 66.7% | | |
| **Tine** | 2010 | 2010 | Bonconto | Senegal | 111.0 | 0 | 23589534 | 44.6% | 0.0% | 0.0% |
| **Traore** | 2021 | 2022 | Bandiagara | Mali | 604.3 | 4.5 | 31315304 | 91.0% | 0.0% | |
| **Zongo** | 2006 | 2007 | Bobo-Dioulasso | Burkina Faso | 0.0 | 0 | 24733476 | 76.9% | 0.0% | NA |
| **Zongo** | 2005 | 2005 | Bobo-Dioulasso | Burkina Faso | 0.0 | 1.5 | 24733476 | 76.9% | 0.0% | NA |
| **Zongo** | 2009 | 2010 | Lena | Burkina Faso | 0.0 | 0 | 25918149 | 75.0% | | |

**Supplementary Table 4 | For each study site and year, a summary of the resistance marker information for SP mutations. For each study site and year, the geographic and temporal distance of the chosen marker survey are chosen, with time difference given in absolute years and distance calculated using the respective longitude and latitude of the trial site and marker survey. The PubMed ID of the chosen surveys are given, along with the prevalence of the three key mutations in the *dhps* gene.**

| Author | Year start | Year end | Location | Country | Distance (km) | PMID | mdr1 86 |
|---|---|---|---|---|---|---|---|
| **Bakyaita** | 2002 | 2003 | Kanungu | Uganda | 29.9 | 16112915 | 89.7% |
| **Bakyaita** | 2002 | 2003 | Kyenjojo | Uganda | 40.5 | 10747279 | 86.0% |
| **Bakyaita** | 2002 | 2003 | Mubende | Uganda | 124.0 | 10747279 | 86.0% |
| **Bell** | 2002 | 2005 | Blantyre | Malawi | 0.0 | 17556619 | 1.8% |
| **Bousema** | 2003 | 2004 | Mbita | Kenya | 34.3 | 22096496 | 81.0% |
| **Clark** | 2005 | 2005 | Kampala | Uganda | 0.0 | 19905933 | 73.0% |
| **Clarke** | 2005 | 2006 | Bondo District | Kenya | 0.0 | 19187521 | 70.4% |
| **Karema** | 2003 | 2004 | All sites | Rwanda | 154.9 | 16112915 | 89.7% |
| **Karema** | 2003 | 2004 | Kicukiro | Rwanda | 376.9 | 21881128 | 92.0% |
| **Karema** | 2003 | 2004 | Mashesha | Rwanda | 514.0 | 21881128 | 92.0% |
| **Karema** | 2003 | 2004 | Rukara | Rwanda | 331.6 | 21881128 | 92.0% |
| **Rwagacondo** | 2001 | 2001 | Kicukiro, Rukara and Mashesha | Rwanda | 154.9 | 16112915 | 89.7% |
| **Menard** | 2004 | 2004 | Moramanga and Saharevo | Madagascar | | 19704124 | 64.3% |
| **Mutabingwa** | 2002 | 2004 | Muheza | Tanzania | 0.0 | 17194834 | 93.2% |
| **Nuwa** | 2022 | 2022 | Karamoja | Uganda | 0.0 | 39826559 | 12.5% |
| **Staedke** | 2002 | 2003 | Kampala | Uganda | 3.9 | 21881128 | 90.6% |
| **Yeka** | 2002 | 2004 | Apac | Uganda | 31.4 | 16112915 | 64.5% |

| | | | | | | | |
|---|---|---|---|---|---|---|---|
| **Yeka** | 2002 | 2004 | Arua | Uganda | 1.8 | 16112915 | 64.6% |
| **Yeka** | 2002 | 2004 | Jinja | Uganda | 2.1 | 16112915 | 77.8% |
| **Yeka** | 2002 | 2004 | Tororo | Uganda | 2.9 | 24799371 | 89.6% |
| **Bojang** | 2007 | 2007 | Upper River Region | The Gambia | 38.7 | 27549635 | 18.2% |
| **Cisse** | 2007 | 2007 | Ndoffane | Senegal | 52.5 | 18008244 | 50.6% |
| **Dicko** | 2008 | 2008 | Kati | Mali | 13.3 | 19245687 | 35.5% |
| **Dicko** | 2008 | 2008 | Djoliba | Mali | 37.5 | 19245687 | 35.5% |
| **Dicko** | 2008 | 2008 | Siby | Mali | 46.6 | 19245687 | 35.5% |
| **Dicko** | 2008 | 2008 | Ouelessebougou | Mali | 73.3 | 19245687 | 35.5% |
| **Djalle** | 2008 | 2009 | Bangui | Central African Republic | 0.0 | 16474071 | 21.9% |
| **Kayentao** | 2005 | 2006 | Faladje | Mali | 64.8 | 19245687 | 35.5% |
| **Konate** | 2008 | 2008 | Toeghin | Burkina Faso | 52.4 | 23153201 | 13.6% |
| **Konate** | 2008 | 2008 | Niou | Burkina Faso | 29.5 | 23153201 | 13.6% |
| **Konate** | 2008 | 2008 | Laye | Burkina Faso | 47.6 | 23153201 | 13.6% |
| **Konate** | 2008 | 2008 | Sao | Burkina Faso | 136.8 | 23153201 | 13.6% |
| **Konate** | 2008 | 2008 | Bousse | Burkina Faso | 33.1 | 23153201 | 13.6% |
| **Maiga** | 2004 | 2005 | Kolle & Bamako | Mali | 1.2 | 19245687 | 35.5% |
| **Mockenhaupt** | 2002 | 2002 | Tamale | Ghana | 1.0 | 16297285 | 56.4% |

| | | | | | | | |
|---|---|---|---|---|---|---|---|
| **Ndiaye** | 2011 | 2011 | Saraya | Senegal | 0.0 | 30865632 | 70.0% |
| **Sesay** | 2008 | 2008 | Farafenni | The Gambia | 144.4 | 27549635 | 17.6% |
| **Sokhna** | 2006 | 2006 | Niakhar | Senegal | 66.4 | 24314037 | 11.6% |
| **Tine** | 2010 | 2010 | Bonconto | Senegal | 60.7 | 27549635 | 17.6% |
| **Traore** | 2021 | 2022 | Bandiagara | Mali | 296.2 | 27662368 | 15.3% |
| **Zongo** | 2006 | 2007 | Bobo-Dioulasso | Burkina Faso | 0.0 | 20231394 | 38.7% |
| **Zongo** | 2005 | 2005 | Bobo-Dioulasso | Burkina Faso | 0.0 | 20231394 | 38.7% |
| **Zongo** | 2009 | 2010 | Lena | Burkina Faso | 9.0 | 24733476 | 29.1% |

**Supplementary Table 5 | For each study site and year, a summary of the resistance marker information for AQ mutations. For each study site and year, the geographic distance of the chosen marker survey are chosen, with time difference given in absolute years and distance calculated using the respective longitude and latitude of the trial site and marker survey. The PubMed ID of the chosen surveys are given, along with the prevalence of the *mdr1* N86Y mutation.**

| *Author* | Start Year | End Year | Location | Country | SP Resistance | AQ Resistance | Classification |
|---|---|---|---|---|---|---|---|
| *Bakyaita* | 2002 | 2003 | Kanungu | Uganda | moderate | high | 4 - moderate SP, high AQ |
| *Bakyaita* | 2002 | 2003 | Kyenjojo | Uganda | moderate | high | 4 - moderate SP, high AQ |
| *Bakyaita* | 2002 | 2003 | Mubende | Uganda | moderate | high | 4 - moderate SP, high AQ |
| *Bell* | 2002 | 2005 | Blantyre | Malawi | moderate | low | 3 - moderate SP, low AQ |
| *Bousema* | 2003 | 2004 | Mbita | Kenya | moderate | high | 4 - moderate SP, high AQ |
| *Clark* | 2005 | 2005 | Kampala | Uganda | moderate | high | 4 - moderate SP, high AQ |
| *Clarke* | 2005 | 2006 | Bondo District | Kenya | moderate | high | 4 - moderate SP, high AQ |
| *Karema* | 2003 | 2004 | All sites | Rwanda | high | high | 6 - high SP, high AQ |
| *Karema* | 2003 | 2004 | Kicukiro | Rwanda | high | high | 6 - high SP, high AQ |

| | | | | | | | |
|---|---|---|---|---|---|---|---|
| *Karema* | 2003 | 2004 | Mashesha | Rwanda | high | high | 6 - high SP, high AQ |
| *Karema* | 2003 | 2004 | Rukara | Rwanda | high | high | 6 - high SP, high AQ |
| *Rwagacondo* | 2001 | 2001 | Kicukiro, Rukara and Mashesha | Rwanda | high | high | 6 - high SP, high AQ |
| *Menard* | 2004 | 2004 | Moramanga and Saharevo | Madagascar | low | high | 2 - low SP, high AQ |
| *Mutabingwa* | 2002 | 2004 | Muheza | Tanzania | low | high | 2 - low SP, high AQ |
| *Nuwa* | 2022 | 2022 | Karamoja | Uganda | moderate | low | 3 - moderate SP, low AQ |
| *Staedke* | 2002 | 2003 | Kampala | Uganda | moderate | high | 4 - moderate SP, high AQ |
| *Yeka* | 2002 | 2004 | Apac | Uganda | moderate | high | 4 - moderate SP, high AQ |
| *Yeka* | 2002 | 2004 | Arua | Uganda | moderate | high | 4 - moderate SP, high AQ |
| *Yeka* | 2002 | 2004 | Jinja | Uganda | moderate | high | 4 - moderate SP, high AQ |

| | | | | | | | |
|---|---|---|---|---|---|---|---|
| *Yeka* | 2002 | 2004 | Tororo | Uganda | moderate | high | 4 - moderate SP, high AQ |
| *Bojang* | 2007 | 2007 | Upper River Region | The Gambia | low | low | 1 - low SP, low AQ |
| *Cisse* | 2007 | 2007 | Ndoffane | Senegal | low | high | 2 - low SP, high AQ |
| *Dicko* | 2008 | 2008 | Kati | Mali | low | low | 1 - low SP, low AQ |
| *Dicko* | 2008 | 2008 | Djoliba | Mali | low | low | 1 - low SP, low AQ |
| *Dicko* | 2008 | 2008 | Siby | Mali | low | low | 1 - low SP, low AQ |
| *Dicko* | 2008 | 2008 | Ouelessebougou | Mali | low | low | 1 - low SP, low AQ |
| *Djalle* | 2008 | 2009 | Bangui | Central African Republic | low | low | 1 - low SP, low AQ |
| *Faye* | 2002 | 2003 | All sites | Senegal | low | low | 1 - low SP, low AQ |

| | | | | | | | |
|---|---|---|---|---|---|---|---|
| *Kayentao* | 2005 | 2006 | Faladje | Mali | low | low | 1 - low SP, low AQ |
| *Konate* | 2008 | 2008 | Toeghin | Burkina Faso | low | low | 1 - low SP, low AQ |
| *Konate* | 2008 | 2008 | Niou | Burkina Faso | low | low | 1 - low SP, low AQ |
| *Konate* | 2008 | 2008 | Laye | Burkina Faso | low | low | 1 - low SP, low AQ |
| *Konate* | 2008 | 2008 | Sao | Burkina Faso | low | low | 1 - low SP, low AQ |
| *Konate* | 2008 | 2008 | Bousse | Burkina Faso | low | low | 1 - low SP, low AQ |
| *Maiga* | 2004 | 2005 | Kolle & Bamako | Mali | low | low | 1 - low SP, low AQ |
| *Mockenhaupt* | 2002 | 2002 | Tamale | Ghana | low | high | 2 - low SP, high AQ |
| *Ndiaye* | 2011 | 2011 | Saraya | Senegal | low | high | 2 - low SP, high AQ |
| *Sesay* | 2008 | 2008 | Farafenni | The Gambia | low | low | 1 - low SP, low AQ |

| | | | | | | | |
|---|---|---|---|---|---|---|---|
| *Sokhna* | 2006 | 2006 | Niakhar | Senegal | low | low | 1 - low SP, low AQ |
| *Tine* | 2010 | 2011 | Bonconto | Senegal | low | low | 1 - low SP, low AQ |
| *Traore* | 2021 | 2022 | Bandiagara | Mali | low | low | 1 - low SP, low AQ |
| *Zongo* | 2006 | 2007 | Bobo-Dioulasso | Burkina Faso | low | low | 1 - low SP, low AQ |
| *Zongo* | 2005 | 2005 | Bobo-Dioulasso | Burkina Faso | low | low | 1 - low SP, low AQ |
| *Zongo* | 2009 | 2010 | Lena | Burkina Faso | low | low | 1 - low SP, low AQ |

**Supplementary Table 6 | A table showing all study sites and years and the respective classifications with respect to SP, AQ and combined resistance classifications as described in Supplementary Table 3.**

### Meta-analyses

#### SP+AQ versus DHA+PPQ

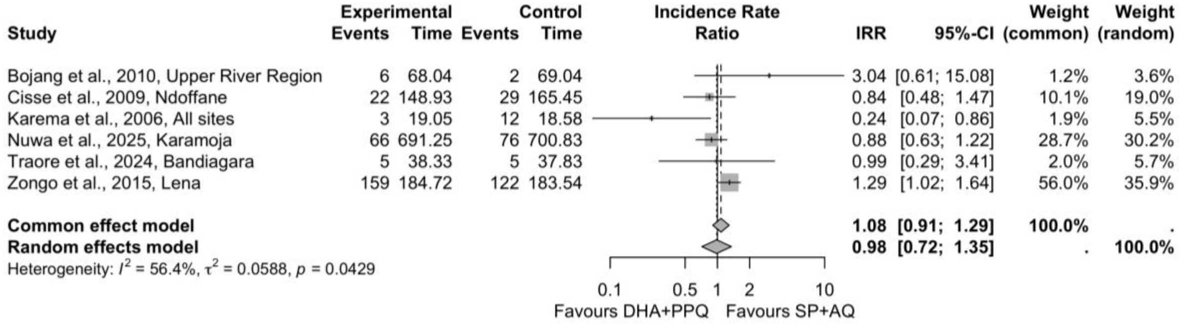

**Supplementary Figure 3 | Meta-analysis comparing DHA+PPQ vs SP+AQ.**

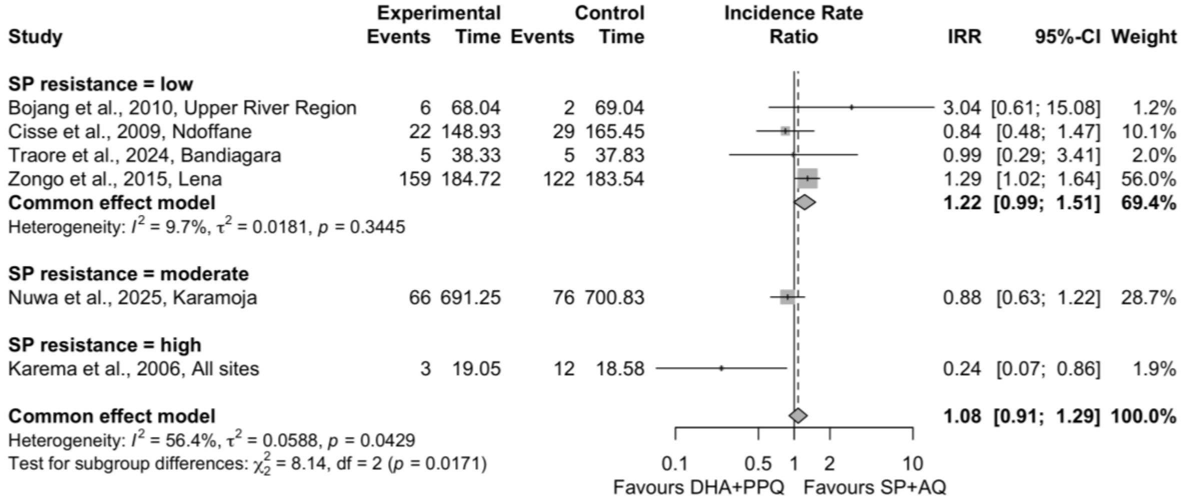

**Supplementary Figure 4 | Meta-analysis comparing DHA+PPQ vs SP+AQ, split by SP resistance classifications as described in Supplementary Table 3**

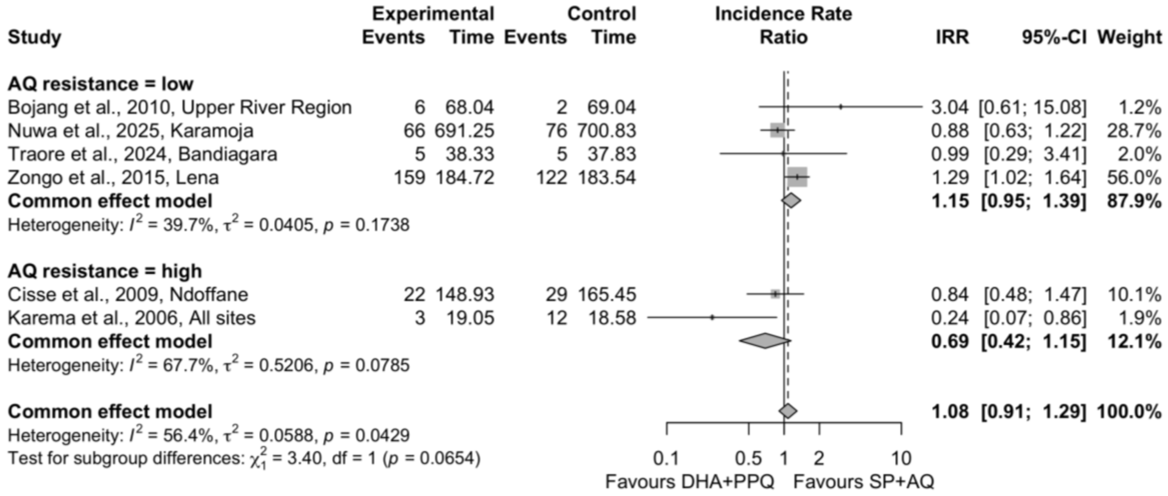

**Supplementary Figure 5 | Meta-analysis comparing DHA+PPQ vs SP+AQ, split by AQ resistance classifications as described in Supplementary Table 3**

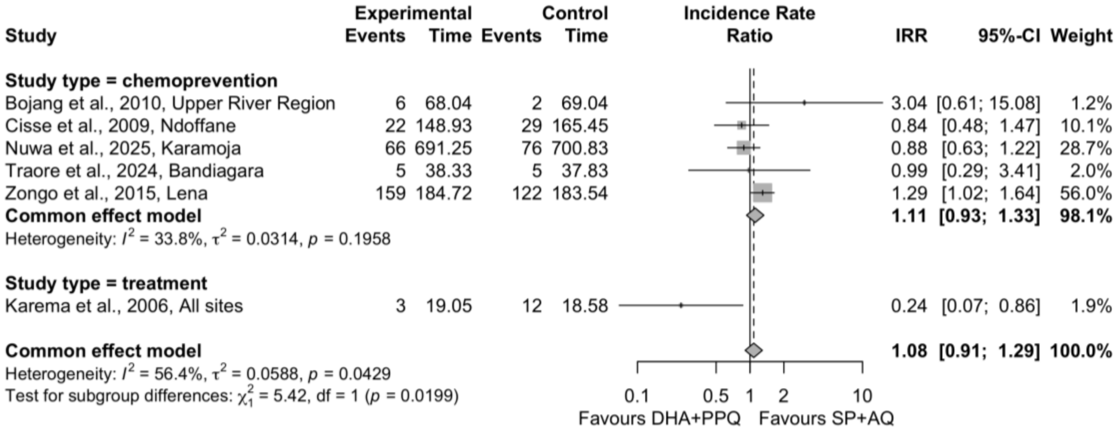

**Supplementary Figure 6 | Meta-analysis comparing DHA+PPQ vs SP+AQ, with subgroup analysis split by the study design.**

#### SP+AQ versus SP/SP+AS

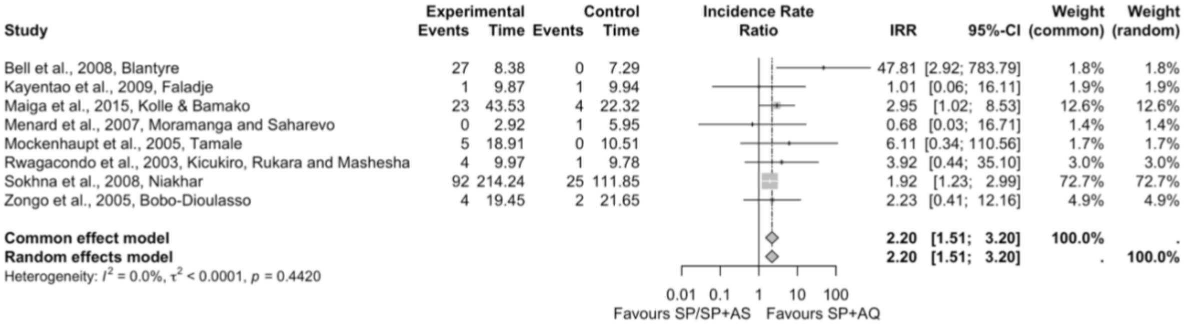

**Supplementary Figure 7 | Meta-analysis of SP+AQ versus SP / SP+AS.**

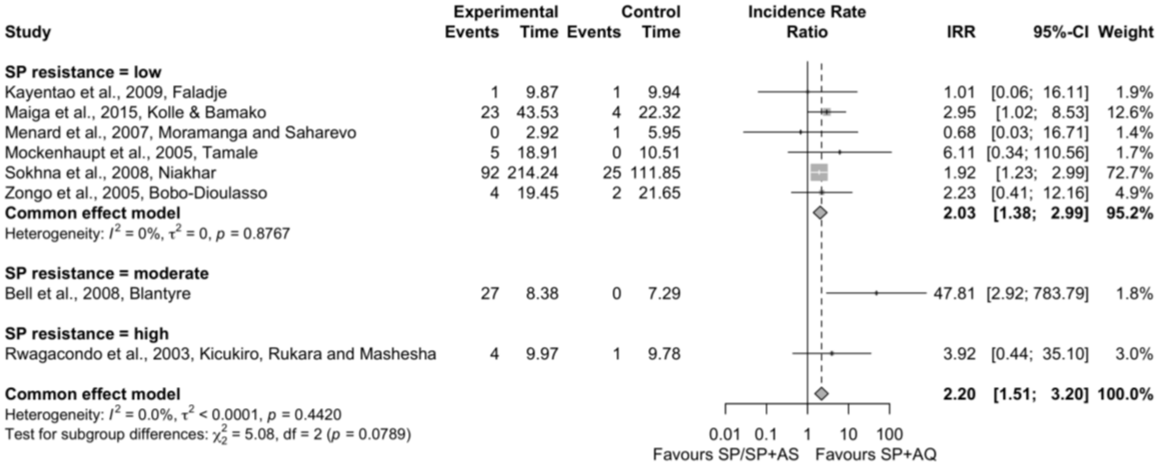

**Supplementary Figure 8 | Meta-analysis comparing SP / SP+AS versus SP+AQ, split by SP resistance classifications as described in Supplementary Table 3**

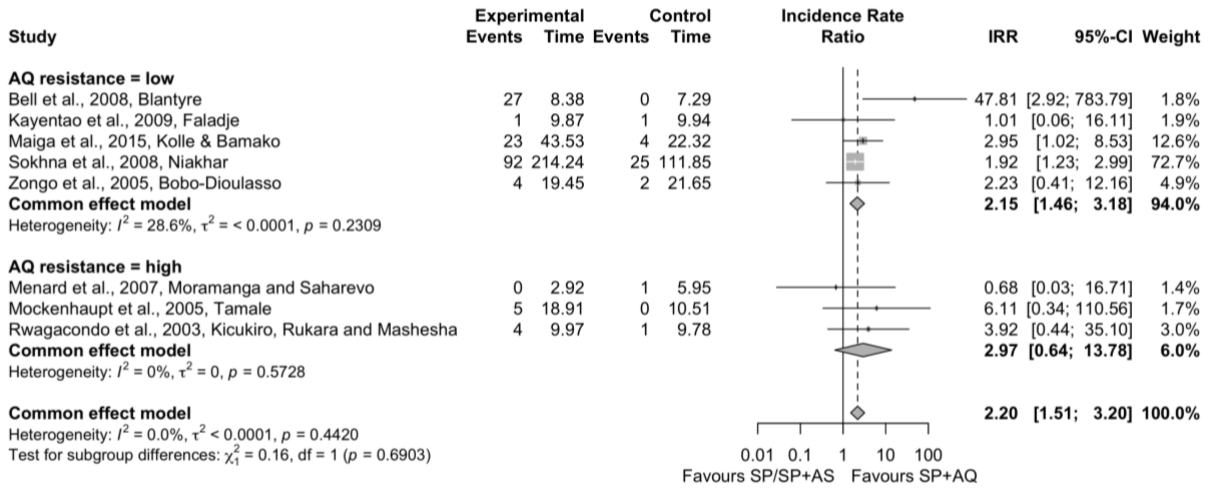

**Supplementary Figure 9 | Meta-analysis comparing SP / SP+AS versus SP+AQ, split by AQ resistance classifications as described in Supplementary Table 3**

### SP+AQ versus AQ/AS+AQ

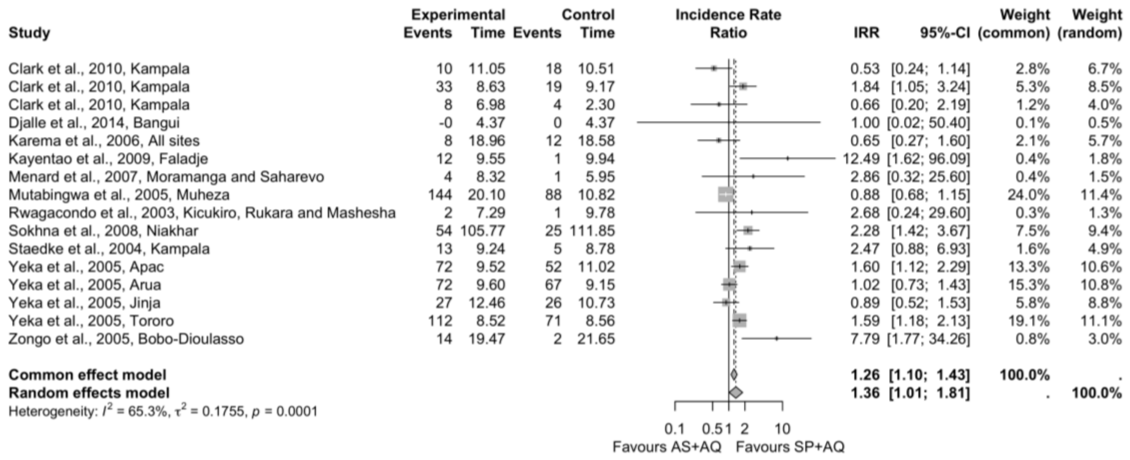

**Supplementary Figure 10 | Meta-analysis comparing AQ / AS+AQ versus SP+AQ**

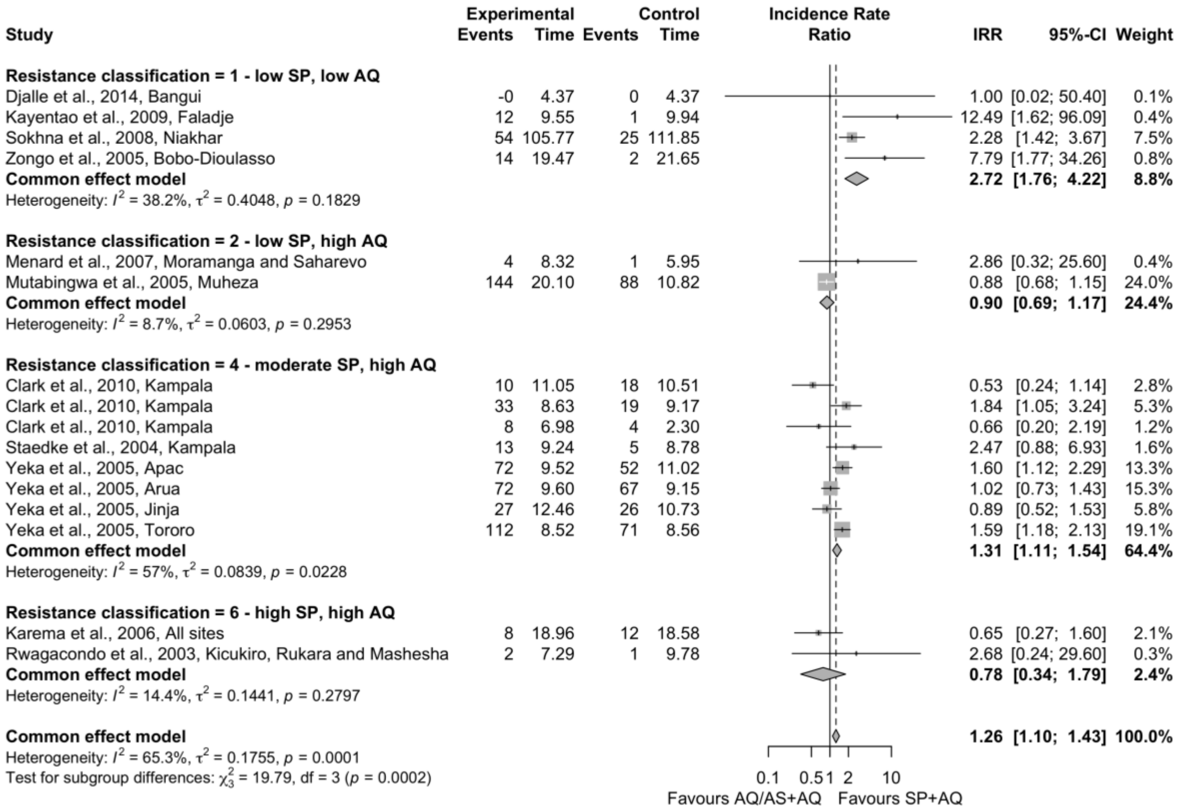

**Supplementary Figure 11 | Meta-analysis comparing AQ / AS+AQ versus SP+AQ, with subgroup analysis by the resistance classification thresholds described in Supplementary Table 3**

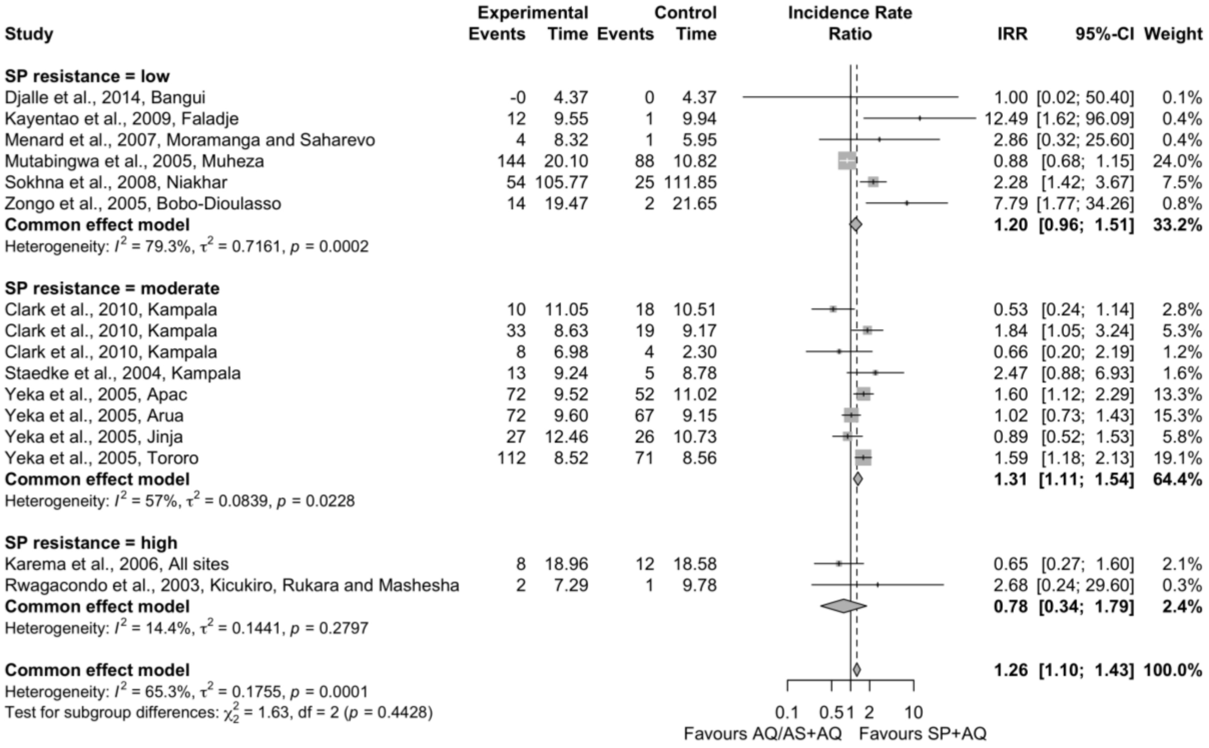

**Supplementary Figure 12 | Meta-analysis comparing AQ / AS+AQ versus SP+AQ, with subgroup analysis by the SP resistance thresholds described in Supplementary Table 3.**

# SP+CQ

| Study | Experimental Events | Experimental Time | Control Events | Control Time | IRR | 95%-CI | Weight (common) | Weight (random) |
|---|---|---|---|---|---|---|---|---|
| Bakyaita et al., 2005, Kanungu | 26 | 2.60 | 26 | 6.36 | 2.45 | [1.42; 4.21] | 7.1% | 7.1% |
| Bakyaita et al., 2005, Kanungu | 4 | 2.02 | 7 | 2.38 | 0.67 | [0.20; 2.30] | 1.4% | 1.4% |
| Bakyaita et al., 2005, Kyenjojo | 32 | 3.63 | 19 | 5.28 | 2.45 | [1.39; 4.33] | 6.5% | 6.5% |
| Bakyaita et al., 2005, Kyenjojo | 21 | 4.22 | 9 | 6.38 | 3.53 | [1.62; 7.70] | 3.4% | 3.4% |
| Bakyaita et al., 2005, Mubende | 47 | 5.05 | 43 | 7.87 | 1.70 | [1.13; 2.57] | 12.2% | 12.2% |
| Bakyaita et al., 2005, Mubende | 12 | 2.90 | 4 | 2.25 | 2.32 | [0.75; 7.20] | 1.6% | 1.6% |
| Bell et al., 2008, Blantyre | 9 | 6.40 | 0 | 7.29 | 21.64 | [1.26; 371.72] | 0.3% | 0.3% |
| Staedke et al., 2004, Kampala | 14 | 5.68 | 5 | 8.78 | 4.33 | [1.56; 12.03] | 2.0% | 2.0% |
| Yeka et al., 2005, Apac | 79 | 8.14 | 52 | 11.02 | 2.06 | [1.45; 2.92] | 17.1% | 17.1% |
| Yeka et al., 2005, Arua | 71 | 5.03 | 67 | 9.15 | 1.93 | [1.38; 2.69] | 18.8% | 18.8% |
| Yeka et al., 2005, Jinja | 37 | 6.15 | 26 | 10.73 | 2.48 | [1.50; 4.10] | 8.3% | 8.3% |
| Yeka et al., 2005, Tororo | 87 | 5.27 | 71 | 8.56 | 1.99 | [1.45; 2.72] | 21.3% | 21.3% |
| **Common effect model** | | | | | **2.10** | **[1.82; 2.43]** | **100.0%** | **.** |
| **Random effects model** | | | | | **2.10** | **[1.82; 2.43]** | **.** | **100.0%** |
| Heterogeneity: $I^2 = 7.7\%$, $\tau^2 < 0.0001$, $p = 0.3698$ | | | | | | | | |

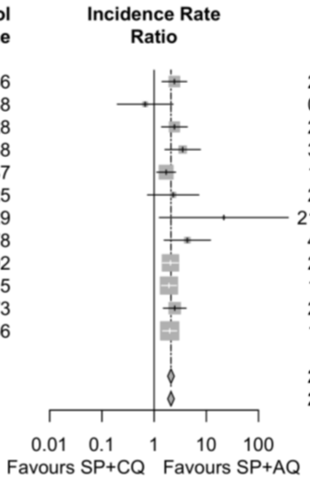

**Supplementary Figure 13 | Meta-analysis comparing SP+CQ versus SP+AQ.**

| Study | Experimental Events | Experimental Time | Control Events | Control Time | IRR | 95%-CI | Weight |
|---|---|---|---|---|---|---|---|
| **SP resistance = moderate** | | | | | | | |
| Bakyaita et al., 2005, Kanungu | 26 | 2.60 | 26 | 6.36 | 2.45 | [1.42; 4.21] | 7.1% |
| Bakyaita et al., 2005, Kanungu | 4 | 2.02 | 7 | 2.38 | 0.67 | [0.20; 2.30] | 1.4% |
| Bakyaita et al., 2005, Kyenjojo | 32 | 3.63 | 19 | 5.28 | 2.45 | [1.39; 4.33] | 6.5% |
| Bakyaita et al., 2005, Kyenjojo | 21 | 4.22 | 9 | 6.38 | 3.53 | [1.62; 7.70] | 3.4% |
| Bakyaita et al., 2005, Mubende | 47 | 5.05 | 43 | 7.87 | 1.70 | [1.13; 2.57] | 12.2% |
| Bakyaita et al., 2005, Mubende | 12 | 2.90 | 4 | 2.25 | 2.32 | [0.75; 7.20] | 1.6% |
| Bell et al., 2008, Blantyre | 9 | 6.40 | 0 | 7.29 | 21.64 | [1.26; 371.72] | 0.3% |
| Staedke et al., 2004, Kampala | 14 | 5.68 | 5 | 8.78 | 4.33 | [1.56; 12.03] | 2.0% |
| Yeka et al., 2005, Apac | 79 | 8.14 | 52 | 11.02 | 2.06 | [1.45; 2.92] | 17.1% |
| Yeka et al., 2005, Arua | 71 | 5.03 | 67 | 9.15 | 1.93 | [1.38; 2.69] | 18.8% |
| Yeka et al., 2005, Jinja | 37 | 6.15 | 26 | 10.73 | 2.48 | [1.50; 4.10] | 8.3% |
| Yeka et al., 2005, Tororo | 87 | 5.27 | 71 | 8.56 | 1.99 | [1.45; 2.72] | 21.3% |
| **Common effect model** | | | | | **2.10** | **[1.82; 2.43]** | **100.0%** |
| Heterogeneity: $I^2 = 7.7\%$, $\tau^2 = < 0.0001$, $p = 0.3698$ | | | | | | | |
| **Common effect model** | | | | | **2.10** | **[1.82; 2.43]** | **100.0%** |
| Heterogeneity: $I^2 = 7.7\%$, $\tau^2 < 0.0001$, $p = 0.3698$ | | | | | | | |
| Test for subgroup differences: $\chi^2_0 = 0.00$, df = 0 ($p$ = NA) | | | | | | | |

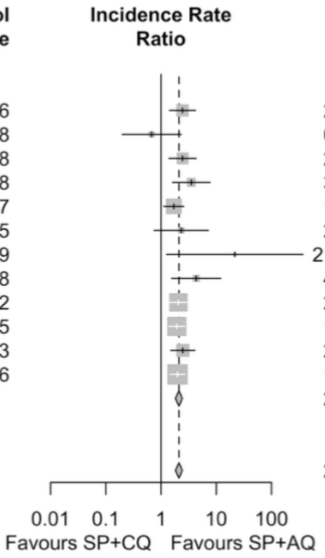

**Supplementary Figure 14 | Meta-analysis comparing SP+CQ versus SP+AQ, split by SP resistance as described in Supplementary Table 3.**

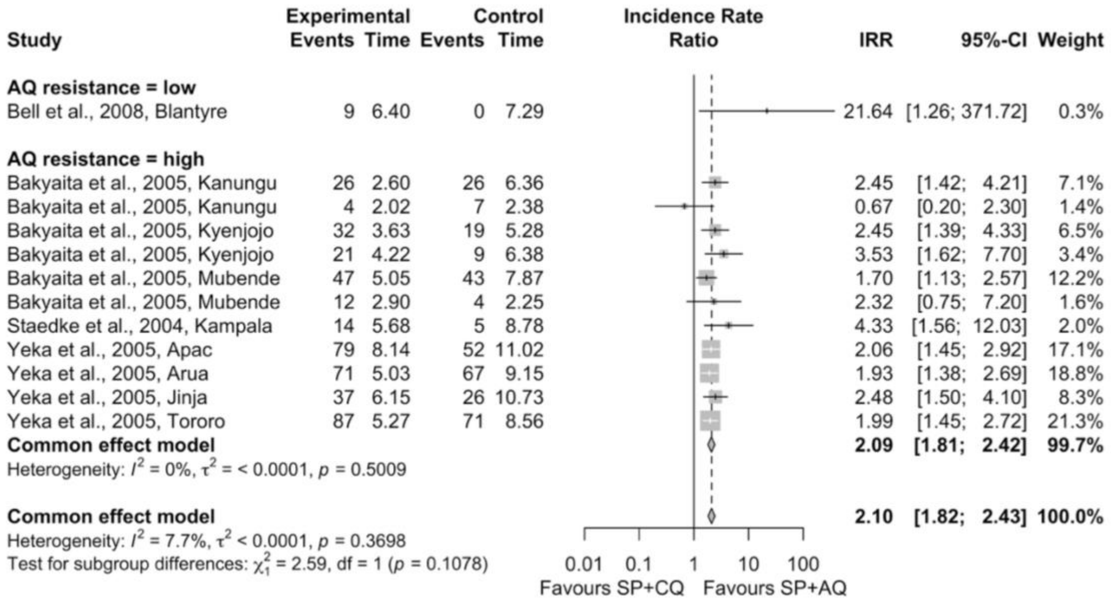

**Supplementary Figure 15 | Meta-analysis comparing SP+CQ versus SP+AQ, split by AQ resistance as described in Supplementary Table 3.**

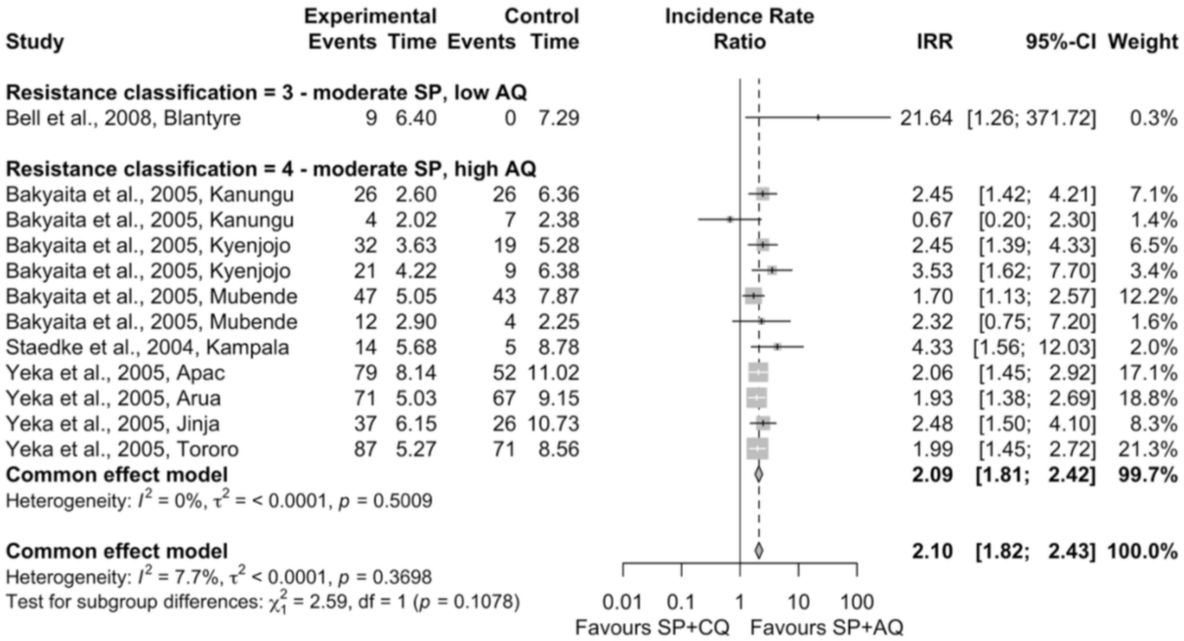

**Supplementary Figure 16 | Meta-analysis comparing SP+CQ versus SP+AQ, split by resistance classification thresholds as described in Supplementary Table 3.**

#### SP+AQ versus control

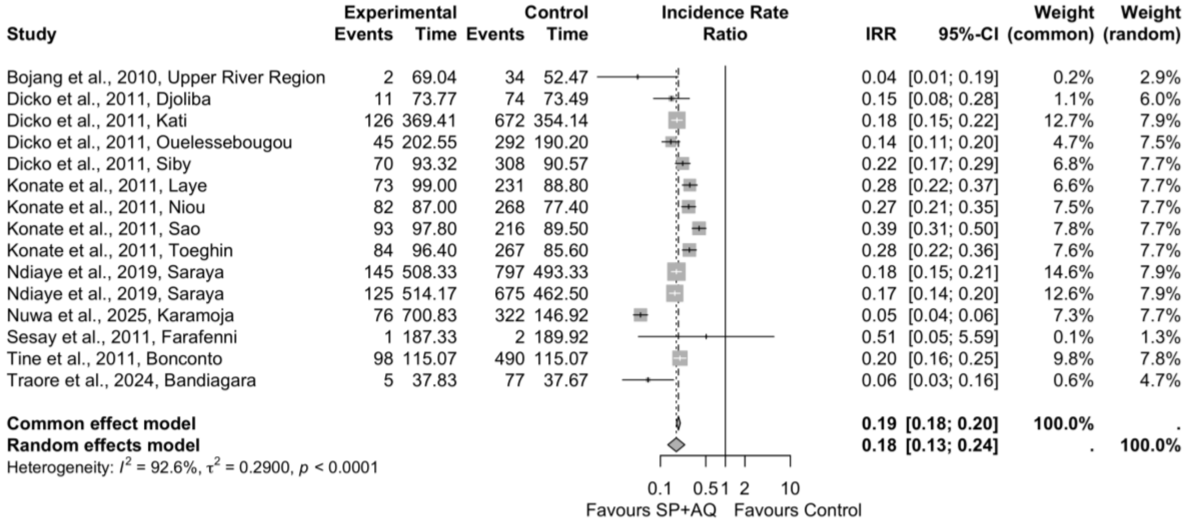

**Supplementary Figure 17 | Meta-analysis of SP+AQ versus control arms.**

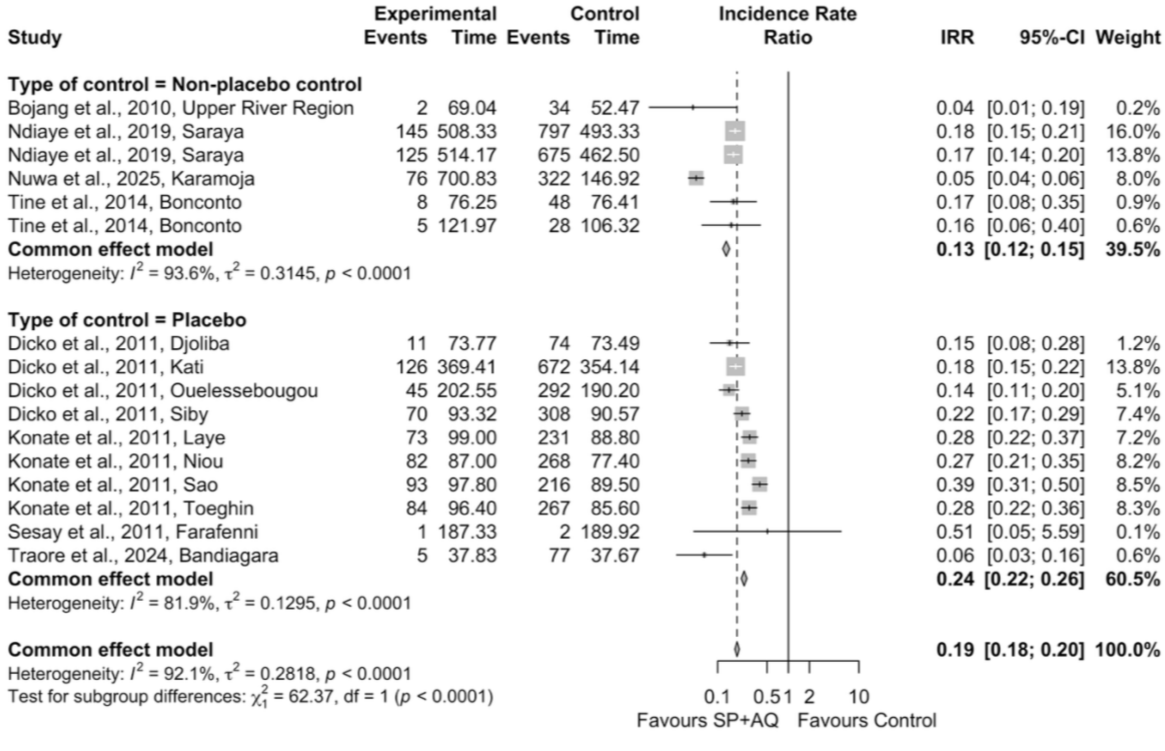

**Supplementary Figure 18 | Meta-analysis of SP+AQ versus control arms, split by trial design.**

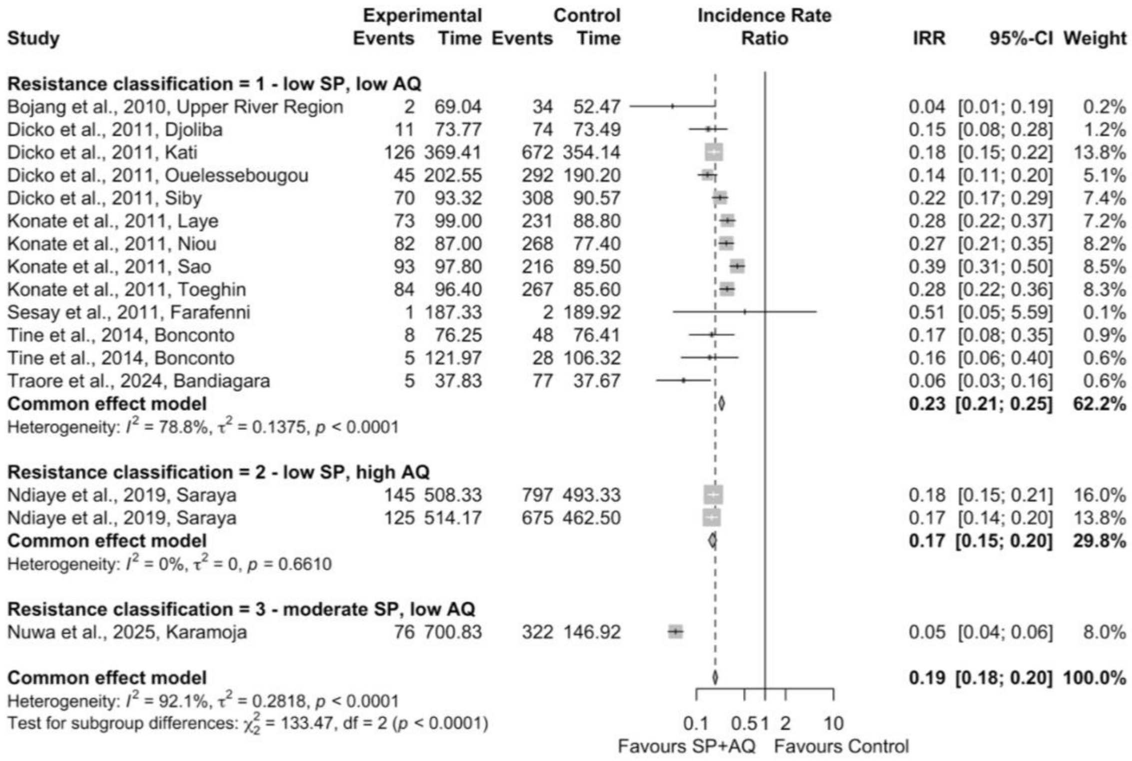

**Supplementary Figure 19 | Meta-analysis of SP+AQ versus control arms, split by resistance classification as described in Supplementary Table 3.**

#### SP+AQ versus SP+PPQ

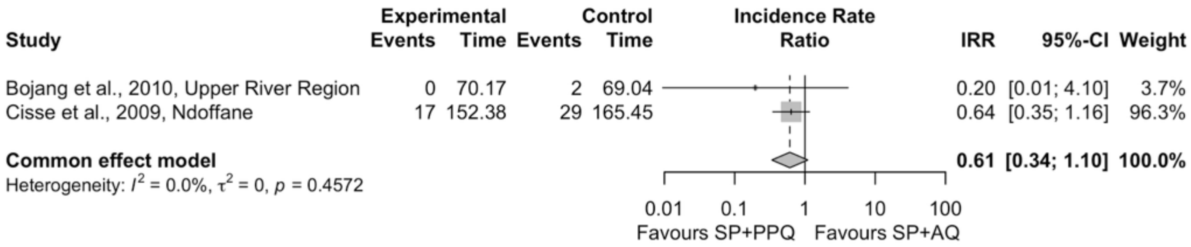

**Supplementary Figure 20 | Meta-analysis of SP+PPQ versus SP+AQ.**

| Section and Topic | Item # | Checklist item | Location where item is reported |
|---|---|---|---|
| **TITLE** | | | |
| Title | 1 | Identify the report as a systematic review. | 1 |
| **ABSTRACT** | | | |
| Abstract | 2 | See the PRISMA 2020 for Abstracts checklist. | |
| **INTRODUCTION** | | | |
| Rationale | 3 | Describe the rationale for the review in the context of existing knowledge. | Introduction |
| Objectives | 4 | Provide an explicit statement of the objective(s) or question(s) the review addresses. | Introduction |
| **METHODS** | | | |
| Eligibility criteria | 5 | Specify the inclusion and exclusion criteria for the review and how studies were grouped for the syntheses. | Methods & SI p5-6 |
| Information sources | 6 | Specify all databases, registers, websites, organisations, reference lists and other sources searched or consulted to identify studies. Specify the date when each source was last searched or consulted. | Methods: search strategy |
| Search strategy | 7 | Present the full search strategies for all databases, registers and websites, including any filters and limits used. | SI p5 |
| Selection process | 8 | Specify the methods used to decide whether a study met the inclusion criteria of the review, including how many reviewers screened each record and each report retrieved, whether they worked independently, and if applicable, details of automation tools used in the process. | Methods: data extraction & QA |
| Data collection process | 9 | Specify the methods used to collect data from reports, including how many reviewers collected data from each report, whether they worked independently, any processes for obtaining or confirming data from study investigators, and if applicable, details of automation tools used in the process. | Methods: data extraction & QA |
| Data items | 10a | List and define all outcomes for which data were sought. Specify whether all results that were compatible with each outcome domain in each study were sought (e.g. for all measures, time points, analyses), and if not, the methods used to decide which results to collect. | Methods |
| | 10b | List and define all other variables for which data were sought (e.g. participant and intervention characteristics, funding sources). Describe any assumptions made about any missing or unclear information. | Methods, SI Methods, data template |
| Study risk of bias assessment | 11 | Specify the methods used to assess risk of bias in the included studies, including details of the tool(s) used, how many reviewers assessed each study and whether they worked independently, and if applicable, details of automation tools used in the process. | SI p9-10 |
| Effect measures | 12 | Specify for each outcome the effect measure(s) (e.g. risk ratio, mean difference) used in the synthesis or presentation of results. | IRR throughout, as stated in all sections |
| Synthesis methods | 13a | Describe the processes used to decide which studies were eligible for each synthesis (e.g. tabulating the study intervention characteristics and comparing against the planned groups for each synthesis (item #5)). | Methods: statistical analysis |
| | 13b | Describe any methods required to prepare the data for presentation or synthesis, such as handling of missing summary statistics, or data conversions. | Methods: |

| Section and Topic | Item # | Checklist item | Location where item is reported |
|---|---|---|---|
| | | | statistical analysis |
| | 13c | Describe any methods used to tabulate or visually display results of individual studies and syntheses. | Methods: statistical analysis |
| | 13d | Describe any methods used to synthesize results and provide a rationale for the choice(s). If meta-analysis was performed, describe the model(s), method(s) to identify the presence and extent of statistical heterogeneity, and software package(s) used. | Methods: statistical analysis |
| | 13e | Describe any methods used to explore possible causes of heterogeneity among study results (e.g. subgroup analysis, meta-regression). | Methods: statistical analysis |
| | 13f | Describe any sensitivity analyses conducted to assess robustness of the synthesized results. | NA |
| Reporting bias assessment | 14 | Describe any methods used to assess risk of bias due to missing results in a synthesis (arising from reporting biases). | SI quality assessment p9-10 |
| Certainty assessment | 15 | Describe any methods used to assess certainty (or confidence) in the body of evidence for an outcome. | Methods: statistical analysis |
| **RESULTS** | | | |
| Study selection | 16a | Describe the results of the search and selection process, from the number of records identified in the search to the number of studies included in the review, ideally using a flow diagram. | Figure 1 |
| | 16b | Cite studies that might appear to meet the inclusion criteria, but which were excluded, and explain why they were excluded. | |
| Study characteristics | 17 | Cite each included study and present its characteristics. | Table 1 |
| Risk of bias in studies | 18 | Present assessments of risk of bias for each included study. | SI |
| Results of individual studies | 19 | For all outcomes, present, for each study: (a) summary statistics for each group (where appropriate) and (b) an effect estimate and its precision (e.g. confidence/credible interval), ideally using structured tables or plots. | Figures in results and SI results. |
| Results of syntheses | 20a | For each synthesis, briefly summarise the characteristics and risk of bias among contributing studies. | |
| | 20b | Present results of all statistical syntheses conducted. If meta-analysis was done, present for each the summary estimate and its precision (e.g. confidence/credible interval) and measures of statistical heterogeneity. If comparing groups, describe the direction of the effect. | Figures in results and SI results. |
| | 20c | Present results of all investigations of possible causes of heterogeneity among study results. | Figures in results and SI results. |
| | 20d | Present results of all sensitivity analyses conducted to assess the robustness of the synthesized results. | NA |

| Section and Topic | Item # | Checklist item | Location where item is reported |
|---|---|---|---|
| Reporting biases | 21 | Present assessments of risk of bias due to missing results (arising from reporting biases) for each synthesis assessed. | NA |
| Certainty of evidence | 22 | Present assessments of certainty (or confidence) in the body of evidence for each outcome assessed. | Figures in results and SI results. |
| **DISCUSSION** | | | |
| Discussion | 23a | Provide a general interpretation of the results in the context of other evidence. | Discussion |
| | 23b | Discuss any limitations of the evidence included in the review. | Discussion |
| | 23c | Discuss any limitations of the review processes used. | Discussion |
| | 23d | Discuss implications of the results for practice, policy, and future research. | Discussion |
| **OTHER INFORMATION** | | | |
| Registration and protocol | 24a | Provide registration information for the review, including register name and registration number, or state that the review was not registered. | Abstract and methods |
| | 24b | Indicate where the review protocol can be accessed, or state that a protocol was not prepared. | Abstract and methods |
| | 24c | Describe and explain any amendments to information provided at registration or in the protocol. | NA |
| Support | 25 | Describe sources of financial or non-financial support for the review, and the role of the funders or sponsors in the review. | Funding declaration |
| Competing interests | 26 | Declare any competing interests of review authors. | Author declarations |
| Availability of data, code and other materials | 27 | Report which of the following are publicly available and where they can be found: template data collection forms; data extracted from included studies; data used for all analyses; analytic code; any other materials used in the review. | All available via GitHub |
